# Effects of Mitigation and Control Policies in Realistic Epidemic Models Accounting for Household Transmission Dynamics

**DOI:** 10.1101/2023.01.26.23285075

**Authors:** Fernando Alarid-Escudero, Jason Andrews, Jeremy D. Goldhaber-Fiebert

## Abstract

**Background:** Compartmental infectious disease (ID) models are often used to evaluate non-pharmaceutical interventions (NPIs) and vaccines. Such models rarely separate within-household and community transmission, potentially introducing biases in situations where multiple transmission routes exist. We formulated an approach that incorporates household structure into ID models, extending the work of House and Keeling.

**Design:** We developed a multi-compartment susceptible-exposed-infectious-recovered-susceptible-vaccinated (MC-SEIRSV) modeling framework, allowing non-exponentially distributed duration in Exposed and Infectious compartments, that tracks within-household and community transmission. We simulated epidemics that varied by community and household transmission rates, waning immunity rate, household size (3 or 5 members), and numbers of Exposed and Infectious compartments (1-3 each). We calibrated otherwise identical models without household structure to the early phase of each parameter combination’s epidemic curve. We compared each model pair in terms of epidemic forecasts and predicted NPI and vaccine impacts on: the timing and magnitude of the epidemic peak and on its total size. Meta-analytic regressions characterized the relationship between household structure inclusion and the size and direction of biases.

**Results:** Otherwise similar models with and without household structure produced equivalent early epidemic curves. However, forecasts from models without household structure were biased. Without intervention, they were upward-biased on peak size and total epidemic size, with biases also depending on the number of Exposed and Infectious compartments. Model-estimated NPI effects of a 60% reduction in community contacts on peak time and size were systematically overestimated without household structure. Biases were smaller with a 20% reduction NPI. Because vaccination impacted both community and household transmission, their biases were smaller.

**Conclusions:** ID models without household structure can produce biased outcomes in settings where within-household and community transmission differ.

## 1 Introduction

Compartmental infectious disease (ID) dynamic transmission models are often used to quantify benefits from interventions intended to mitigate and control respiratory infectious diseases. However, they may fail to provide accurate forecasts or estimates of intervention effects due to simplifications they necessarily make regarding mixing and transmission patterns, rarely separating transmission within households from community transmission. Empirical evidence from a range of respiratory infectious diseases suggests that within-household transmission can be substantially more intense than community transmission[Fun+20; Mad+22; Dah+21; Gly+18]. While dynamic transmission microsimulation models with explicit contact networks avoid the need to make this simplification, they can be computationally intensive, especially when modeling large populations, and extensive data are required to parameterize their contact networks.

As a feasible alternative, past work developed approaches for explicitly incorporating household transmission into compartmental Susceptible-Infected-Recovered (SIR) models [HK08]. In SIR models, household size and transmission affect the dynamics of outbreaks, including infection incidence, with the time course and level of the overall epidemic depending on how household and community transmission differ and on household sizes [HK09]. Furthermore, the effects of non-pharmaceutical interventions (NPIs) predicted with a SIR model that does not separately model household transmission can differ if, for example, an NPI (e.g., business closures) reduced community contacts differently than household contacts as was shown using mobility and collocation data early in the COVID-19 pandemic[Goo23; Dat23; Dat22].

The utility of this past work could be substantially increased if the approach were extended to more complex compartmental models. Sometimes it is necessary for realism to implement multi-compartment Susceptible-Exposed-Infected-Recovered (SEIR) models of open populations and to include symptomatic infections, case detection, venue-specific transmission, and combinations of NPIs and vaccination [KR08]. For example, when the Mexico City Metropolitan Area (MCMA) faced risks of COVID-19 outbreaks, using a network microsimulation model of its population of almost 20 million would not have been feasible, but the compartmental model used for this epidemic required many of these characteristics [Ala+21].

Our study extends the mathematical framework for the household and community transmission model based on House and Keeling’s work to include more complex compartmental models and a suite of interventions. We illustrate the relevance of these extensions by showing how failure to incorporate household transmission into more complex compartmental models in settings where community and household transmission differ can bias epidemic predictions and intervention effect estimates. Further, we characterize the systematic ways this exclusion biases such outcomes and depends on household size in conjunction with other model features. While all models are wrong in the sense that they simplify reality, decision-analytic models are most useful when they capture dynamics relevant to the decisions that they are used to consider; hence, we are not arguing that explicit modeling of household structure is always required but rather that it is often needed and useful when community and household transmission differ. To this end, we provide an open-source implementation of this extension, facilitating its use by other analysts and incorporation into their models.

## 2 Mathematical Framework for Community and Household Models

### 2.1 SIR to MC-SEIRSV with Interventions

The epidemiology of many IDs can be described as a multi-compartment susceptible-exposed-infected-recovered-susceptible-vaccinated (MC-SEIRSV) model with demography [KR08]. In the model, exposed (*E*) compartments represent individuals that are infected but not yet infectious, and infectious (*I*) compartments represent individuals that are both infected and infectious (i.e., can infect susceptibles (*S*) if contact occurs) [KR08]. By using multiple levels for the *E* and *I* compartments and allowing for differential rates of symptom onset and detection (described below), the model can represent asymptomatic infectiousness and varying infectiousness over the course of infection. Figure 1 depicts the generalized structure of the MC-SEIRSV compartmental model. Each compartment represents part of the population characterized by their disease, diagnostic, and vaccination status.

**Figure 1:**
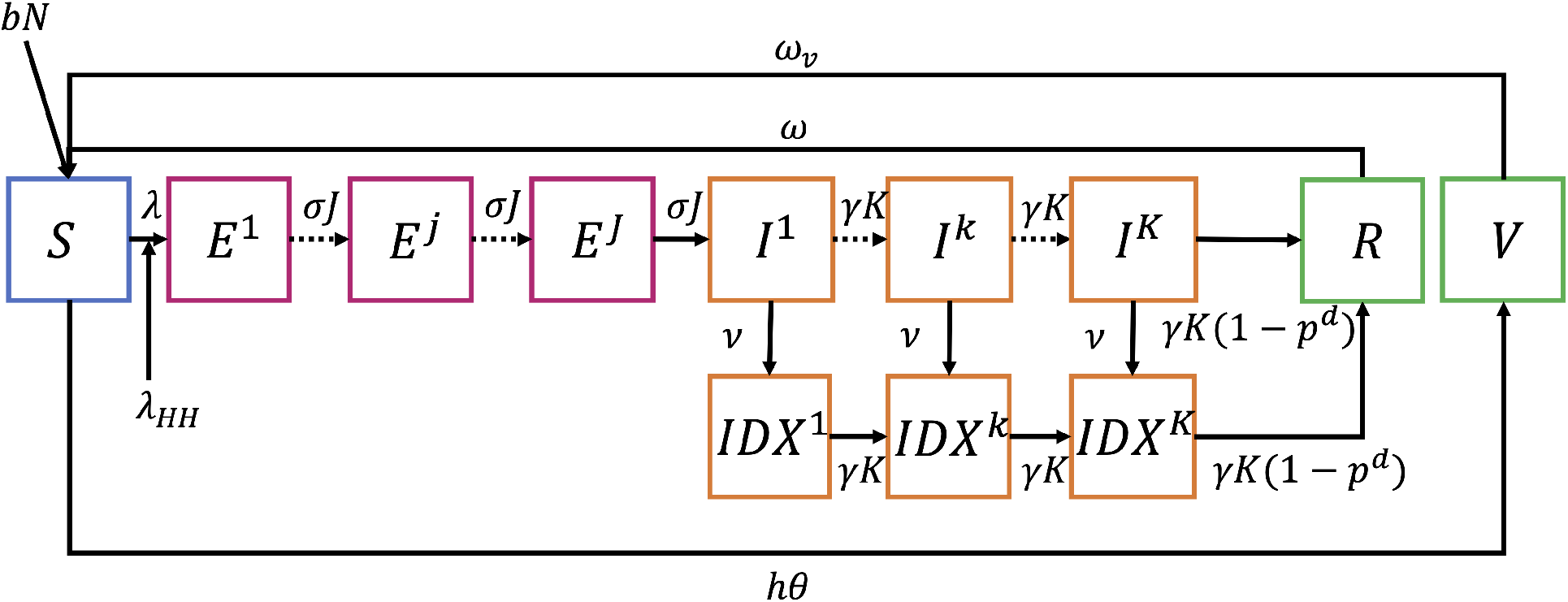
Generalized model diagram. Not shown in the figure is death from other causes at a rate (*μ*) which applies as an outflow from all compartments.

The full model encompasses multiple disease, demographic, and intervention processes. People are born into the susceptible compartment *S* at a rate *b* and face a background mortality rate (*μ*) from all compartments. People can become infected at a rate *λ* and enter the exposed compartment, E, progressing to becoming infectious and moving to *I* before recovering and entering *R*. We stratify the infectious compartments by people’s diagnostic status (*DX*). Infected people face an excess risk of death from their infections (*p*^*d*^), which may be reduced by treatment. Those who recover may lose their immunity at a rate *ω* and become susceptible. We ensure realistic distributions of times in *E* and *I* by using a multi-compartment structure, employing multiple *E* and *I* compartments and rates of progression (*σ*) from *E* to *I* and (*γ)* from *I* to *R* that are multiplied by the number of each type of compartment (*J* and *K*, respectively) [KR08]. Health states with a multi-compartment structure have non-exponentially distributed dwell times (NEDT) following an Erlang distribution. If there is only a single compartment, dwell times are exponentially distributed (EDT) [KR08]. The total size of the population at time *t* is *Pop*, the sum across all compartments and stratifications.

To capture the dynamics of many infectious disease epidemics, including COVID-19, it is important to consider both community and household transmission [Pel+20]. We term the model’s components described until now the “community submodel” (i.e., the non-household components) to differentiate them from the “household submodel”, described below.

In brief, the household submodel acknowledges that people are generally embedded within households, implying that: 1) once a given household’s members are all infected and/or recovered, no further transmission occurs within that household without waning or births or other household entries; and 2) if households are not entirely isolated from one another such that community transmission is still occurring, the interaction between the susceptible and infectious individuals within the household can drive additional community transmission via the within-household force of infection, *λ*_*HH*_(*t*), (related to the household secondary attack rate, e.g., [Fun+20]) a component of the overall force of infection. These household embeddings are also essential when interventions (e.g., shelter-in-place) differentially impact community transmission and household transmission. By extending the prior approach in [HK08] and combining it with our community submodel, we produce an overall population model.

To describe overall transmission dynamics in the population, we first describe transmission in the community submodel, then the key elements of the household submodel, and how the household submodel’s transmission is integrated into the community submodel.

### 2.2 Community submodel with transmission from the household force of infection

The community MC-SEIRSV model consists of a system of ordinary differential equations (ODEs) (see Appendix A.1.1).

#### 2.2.1 Force of infection

The force of infection (FOI) governs the transmission of infection within a population, defined as the instantaneous per-capita rate at which susceptibles acquire infection. FOI reflects the degree of contact between susceptibles and infectious individuals and the pathogen’s per-contact transmissibility. Because contacts can occur in the community and within a household, we construct community- and household-specific FOIs. The community FOI, *λ*(*t*), represents the rate of disease transmission from infectious people in the community (see Appendix A.1.2); the household FOI, *λ*_*HH*_(*t*), is described below; and the linkage between the household and community submodels in terms of FOIs is described in pseudo-code in A.1.3 in the appendix.

#### 2.2.2 Interventions impacting the force of infection

The model incorporates two epidemic control interventions: NPIs and vaccination. NPIs reduce the community FOI by a value *ϕ* ∈ [0, 1]. Susceptible individuals are vaccinated at a rate *h* with a proportion, *θ* ∈ [0, 1], effectively immunized. Successfully vaccinated individuals cannot be infected until their immunity wanes at a rate *ω*_*v*_.

### 2.3 Household submodel

As with [HK08], the household submodel tracks the proportion of households whose members are in various states of the disease’s natural history. For example, in a given population at a given time, 5% of all 3-person households might have 1 member susceptible and 2 members recovered (*HH*_(*S*=1,…,*R*=2)_). We denote the proportion of households whose members are in any combination of community MC-SEIRSV model states (i.e., counts of members in each state which we abbreviate *sc* for state counts) as *HH*_*sc*_, where ∑_*sc*_ *HH*_*sc*_ = 1.

The number of distinct *HH*_*sc*_’s representing households with different state counts grows with household size (*hhsize*) and the number of states in the community submodel (*states*). The equation for the number of household proportions (and hence the number of differential equations in the household sub-model) with a fixed household size is:

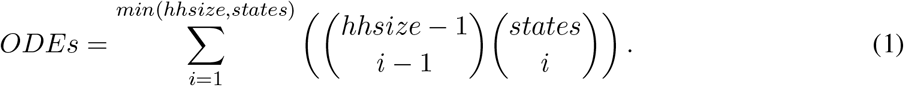

The above equation is based on a sum of products of combinations of partitions. The first combination operator in the equation gives number of unique partitions of a given household size (*hhsize*) into a number of subgroups (*i*). The second combination operator gives the unique number of sets of MC-SEIRSV states (*states*) into which the partitioned subgroups of household members (*i*) can be placed. The reason that the sum is over the minimum of the *hhsize* and *states* is that we cannot partition household members into more subgroups than there are different states into which to place them (*states* < *hhsize*) nor can we assign fewer than 1 household member to a given state (*states* > *hhsize*). Finally, we note that the combination operator where we want “X choose 0” is equal to 1 by definition. As an example, if *hhsize* were 4 and the model was SIR (*states* = 3), then we could have 3 unique ways in which all 4 household members were in 1 state, 3 unique ways in which 3 household members were in 1 state and 1 household member was in another state, and 3 unique ways where 2 household members were in 1 state and 1 household member was in each of the 2 remaining states in the model.

To keep the number of household types and differential equations manageable, we assume that all households are the same size as the average household for a given population, rounded to the nearest whole integer.

We ensure that the household and community submodels’ initial states are consistent. For a given total population size at the start of the community submodel *Pop*_*t*=0_ under the assumption of all households being size *hhsize* = *hhsize*_*avg*_, the number of households is *N*_*households*_ = *Pop*_*t*=0_/*hhsize*_*avg*_. If there is one person in the *E*_1_ state in the entire population, then the fraction of households that have an infected member is 1/*N*_*households*_ and the remainder are households with all susceptibles: (*N*_*households*_ − 1)/*N*_*households*_. This initialization generalizes to more than one starting exposed, infectious, or recovered individual easily under the assumption that the households in which the initial few infections occur are not correlated (i.e., if there are 3 infections, they occur in 3 separate households because *N*_*households*_ is much larger than the initial number of infections). This assumption can be relaxed without loss of generality.

The household submodel’s dynamics include: progression, recovery, waning immunity, vaccination, within-household and community-household transmission, births, and deaths. Modeling many of these dynamics in the household submodel is somewhat more complicated than in the community submodel because the household submodel tracks the fraction of households in a set of discrete states characterized by counts of members in each community MC-SEIRSV model state.

The following example provides the intuition of how the household submodel handles progression and recovery. To simplify the exposition, we ignore the multi-compartment nature of the exposed and infectious states and consider only progression. If there are 4 household members (1 susceptible, 3 exposed, 0 infectious) at a given time, then it is possible that 0, 1, 2, or all 3 of the exposed members will progress to infectious on a given day. Hence, the possible states that this household could be in on the next day include either (1 susceptible, 3 exposed, 0 infectious); (1 susceptible, 2 exposed, 1 infectious); (1 susceptible, 1 exposed, 2 infectious); or (1 susceptible, 0 exposed, 3 infectious). In this example, the frequency of households moving to each state follows a binomial distribution with the probability related to the rate of progression *(σ*). When considering multiple exposed and infectious compartments in the community MCSEIRSV model, the binomial distribution’s probability is then related to *σJ*. Recovery follows a similar pattern where the frequencies of resulting household states follow a binomial distribution with probability related to *γ* (in the multi-compartment model, *γK*). Waning immunity (*ω* > 0) implies that the frequency of household members in *R* moving back to *S* also follows a binomial distribution.

Models of counts of household members where *E* and/or *I* are multi-compartment are, in fact, more complex, with the counts of household members that progress and/or recover following a convolution of binomial distributions. Consider the following example. For each source state (e.g., *E*_2_) which at time *t* might have *h* household members in it, 0, 1, …, *h* members may progress to *E*_3_ at time *t* +1 with the rest remaining in *E*_2_. The counts of progressors are binomially distributed. However, if there are also some household members in *E*_3_ at time t, then the count of people in *E*_3_ at time *t*+1 is more complicated because it depends on the count of the progressors from *E*_2_ as well as the count of the non-progressors among those in *E*_3_. Hence, we arrive at a convolution of binomial distributions in equation (10) with a simple binomial distribution for cases where individuals are in one source state and none in the destination state at time *t*. The formal construction of the convolution of binomial distributions is shown in Appendix A.1.5.

#### 2.3.1 Household force of infection

There are both within-household and community-household transmission routes in the household submodel. Within-household transmission involves infectious household members infecting susceptible household members. Community-household transmission involves infectious individuals in the community (i.e., no non-household members) infecting susceptible household members.

Within-household transmission is related to a) the current number of infectious household members; 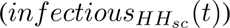; b) the rate of contact between household members; c) the probability of within-household transmission given household contacts (*τ*). The number of infectious individuals in the household is given directly by *HH*_*sc*_. The number of implied daily household contacts for a particular jurisdiction can be computed from household mixing matrices such as those published by Mossong et al., [Mos+08]. Finally, note that because the intensity of household contacts may differ from contacts in the community, the probability of transmission conditional on household contacts (*τ*) can differ from the probability of transmission given community contacts (*β*).

The household FOI, *λ*_*HH*_(*t*), is defined separately from the community FOI, *λ*(t), and connects the dynamics of the household submodel back to the community submodel. *λ*_*HH*_(*t*) depends on the number of within-household contacts between susceptible and infectious members, *contacts*_*HH*_, and the probability of transmission given a household contact (*τ*). In the community submodel, when infectious individuals are detected, their probability of transmission, *β*, is reduced by a factor *f* ∈ [0, 1]. So too, in the household submodel, *τ* is scaled by the same constant yielding *τ′*. However, a necessary simplification for scaling *τ* to produce *τ′* is that the fraction of household infectious contacts to which this scaling factor applies is assumed to be the same as the fraction of prevalent infectious individuals who are currently detected in the community submodel. Therefore, we define the household FOI as

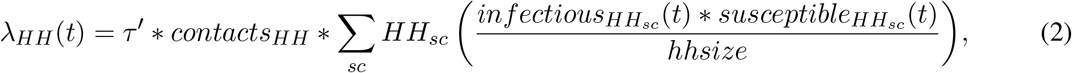

which is the weighted average of within-household transmission (higher where there are more infectious and susceptible individuals simultaneously present in the household), where the weight is the fraction of households with these counts of infectious and susceptible members. The rate of infections generated by household transmission, *λ*_*HH*_(*t*), increases the flow of new infections from *S* to *E* as tracked in the community submodel.

## 3 Simulations and analyses

To illustrate the value of our community-household modeling framework, we analyze its predictions compared to models that do not differentiate household and community transmission. Specifically, we perform analyses to assess how not including household transmission in more complex compartmental models can alter both predictions of epidemic outcomes and intervention effects. Outcomes include the cumulative epidemic magnitude along with the timing and height of epidemic peaks and how these are changed through interventions. As we are interested in situations where household and community transmission differs, we characterize systematic relationships between the size and direction of outcome biases and the population’s average household size in conjunction with other model features and parameters. We employ a design-of-experiments approach, simulating outcomes across a range of household sizes, numbers of exposed and infectious compartments, transmission and recovery patterns, and in the presence/absence of various interventions. We use meta-analytic regressions to characterize the patterns of bias across the model simulations.

### 3.1 Interventions

We employ a set of stylized interventions to carry out our analyses. We consider higher and lower levels of contact reductions via NPIs and different levels of vaccine coverage and vaccine effectiveness. NPI effectiveness (*ϕ*) reduces community contacts by 20% or 60%. Vaccine coverage (*h*) is 30% or 90%, and vaccine effectiveness (*θ*) is 50% or 90%. As public health responses take some time, we initiate interventions 10 days after the model’s start time, roughly at a point where the 50% interquantile range of the number of detected cases is between 8 and 115 infections per 100,000 across the different combinations of model parameter values in the absence of interventions.

### 3.2 Outcomes

We focus on three model-generated outcomes, O: epidemic size, defined as the integral of the prevalent infection epidemic curve; epidemic peak, defined as the maximum number of prevalent infections on a given day; and time when the epidemic peak occurs (see Appendix A.1.6).

### 3.3 Control measures’ effectiveness

We analyze the outcomes and the effects of control measures on them. We define each control measure’s effectiveness (Δ*O*) as the difference in outcome with no control measure (*O*^*nc*^) and the outcome with the control measure (*O*^*c*^), given by

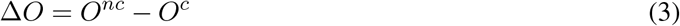

Control measures generally reduce the number of cases; hence, Δ*O* will tend to be a larger positive quantity for more effective measures. Control measures also generally delay the epidemic peak, so for this outcome, Δ*O* will tend to be a larger negative quantity for more effective measures.

### 3.4 Calibration for bias assessment

We evaluate how failure to include household size and other features in the model produces a bias in model outcomes and estimated control measure effects. We sought to enable an interpretable comparison of otherwise similar models that only differ by their inclusions/exclusion of household transmission. Specifically, we imagined an analyst in the early days of an epidemic deciding whether to include household transmission in a dynamic model for epidemic forecasting and considering interventions and their potential effects. If the truth is that household transmission occurs and a household structure should be included, how much and what kinds of biases occur in modeled outcomes if the household transmission is omitted?

Hence, for each combination of the number of exposed compartments, number of infectious compartments, household sizes greater than 1 (i.e., models with household structure), household transmission rate, community rate, and waning rate in Table A1, we generate an epidemic trace of incident daily infections for the first 15 days of the epidemic in the absence of interventions. For each combination, we then instantiate a corresponding model with the same parameters except that it had no household transmission (*τ* = 0 and household size of 1) and calibrate its community transmission parameter (*β*) such that the model without household transmission produces daily incident infections in the first 15 days that match the corresponding model with the household transmission. Since we consider a deterministic case without uncertainty, we focus on the point estimate of *β*, yielding an incidence that matches the model with the household transmission. We used the Nelder-Mead algorithm for calibration, minimizing the sum-of-squared errors between the models’ daily incidence (with household transmission vs. no household transmission).

We use the corresponding pairs of models (those with and without household transmission that have the same epidemic trace for the first 15 days) to assess how predicted outcomes and intervention effects might be biased. We quantify the bias as the absolute or percentage change in a given control measure’s effectiveness as predicted with a model that includes both community and household transmission (*HH*) versus as predicted with the corresponding calibrated model that only includes community transmission (*NH*). The absolute bias (*aBias*) is defined as

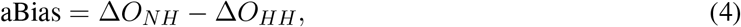

If the effect of an intervention is a reduction in an outcome (e.g., total infection days, peak infections, etc.), then an absolute positive bias implies that the estimated effectiveness of control measures is smaller when the household transmission is not included (i.e., underestimated effectiveness). If the effect of an intervention is to increase an outcome (e.g., life expectancy, time to epidemic peak, etc.), then an absolute positive bias implies that the estimated effectiveness is larger when the household transmission is not included (overestimated effectiveness).

### 3.5 Design of experiments

We use a meta-analytic regression approach relating epidemic outcomes and biases to household size, numbers of exposed and infectious compartments, and community and household transmission rates. The regressions also include two-way interactions between household size and the number of exposed and infectious compartments, allowing for non-linear relationships.

For epidemic outcomes in the absence of intervention, we estimate the regressions for each outcome based on a set of parameter values generated following a full factorial design of experiments (DoE) (Table A1).[HKP17; MWC19]

We use the same general approach (i.e., DoE design and meta-regression) for intervention effects. We focus on the intervention effects of NPIs that would be differentially delivered in communities vs. households (e.g., business closures) without vaccination. In supplementary analyses, we consider vaccination in the absence of NPIs.

## 4 Results

### 4.1 Household structure and calibrated model parameter values

Otherwise, similar models with and without household structure can produce equivalent early epidemic curves across a broad range of disease parameters. However, this does not guarantee that their longer-term epidemic projections or projected intervention effects will be similar. With otherwise similar modeling choices at the beginning of an outbreak, models that do not include household structure calibrated to the initial period’s rise in infection prevalence correspond closely to otherwise similar models with a household structure for our study’s 648 natural history parameter combinations. Calibrations of all models lacking household structure converged, producing epidemic curves whose daily prevalent infections differed from corresponding models with household structure by 1% on average and all ≤ 2.25%. For all parameter combinations, calibrated models without household transmission had higher community transmission rates (*β*) than corresponding models that include a household structure to offset household transmission (*τ*) that they did not include. On average, *β* in the models without household structure was 0.23 higher than in corresponding models with household structure. Meta-analytic regressions revealed that calibrated /3s for models without household structure were significantly higher when either *β* or *τ* in the corresponding model with a household structure was higher, and even more so for models with larger household sizes and in models with more *E* compartments (Appendix Table A2).

### 4.2 Impact on model-predicted, longer-term natural history

Failure to include household structure can cause modeled longer-term epidemic natural history to differ from otherwise similar models across a broad range of disease parameters. Figure 2 shows the natural history epidemic curves for an exemplar parameter set for which we systematically varied the number of exposed and infectious states. For this example, with household size=3, when the household structure is not included, the model’s epidemic curve peak is higher, earlier, and drops sooner than the model that includes household transmission. The differences appear slightly larger when there are multiple *E* compartments. More generally, Appendix Tables A3 - A4 show that failure to include household structure when projecting the epidemic natural history results in higher peak infections on average with slightly higher variation across parameters sets and also slightly higher epidemic size with slightly lower variance.

**Figure 2:**
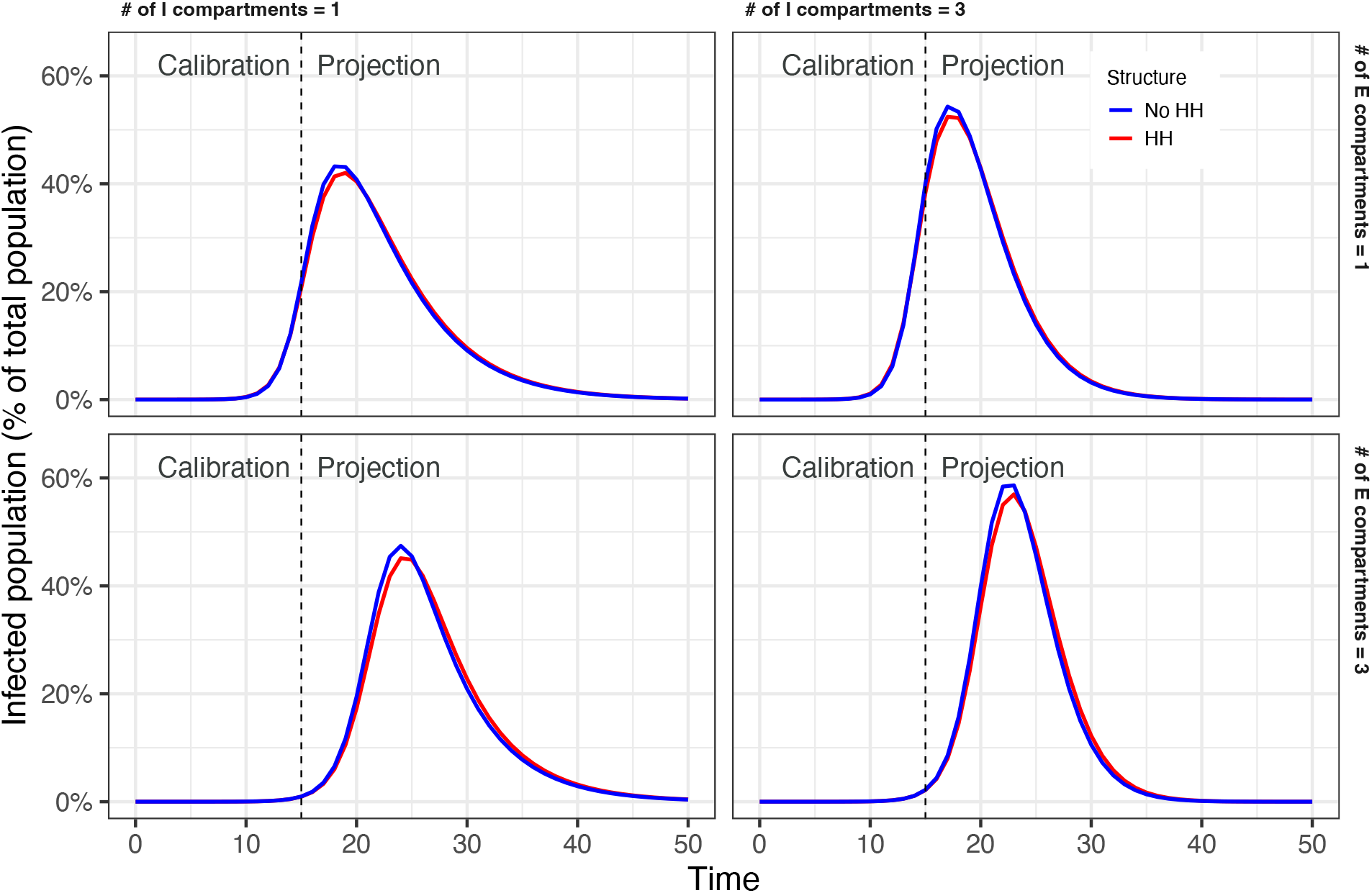
Natural history epidemic curves.

Using meta-analytic regressions to examine outcomes without intervention, we found that, on average, excluding household structure did not significantly affect peak time. However, the trend was towards having slightly later peaks with larger household sizes. The epidemic peak was approximately 25,000 people larger (5% larger) than the epidemic peak size when the household structure was excluded for a setting with a true household size of 3 or 5. The total epidemic size over 100 days was approximately 60,000 larger (1.2% larger) than the total epidemic size when the household structure was excluded, and the true household size was 3 (approximately 57,000 larger (1.1% larger) if the true household size was 5) (Table 1). The magnitude of the differences, particularly for peak size, varied slightly depending on the number of exposed and infectious states; the more *E* and *I* compartments, the greater the overestimate of the peak in an otherwise similar model without household structure compared to one with household structure.

**Table 1:** Meta-regression estimates on absolute differences in outcomes of excluding household structure in the absence of intervention.

|  | Peak time (days) |  | Peak size (people) |  | Epidemic size (people) |  |
| --- | --- | --- | --- | --- | --- | --- |
| Household size 3 (HH3) | -0.3* | -0.3 | 25,646.6*** | 24,603.8*** | 136,783.1*** | 139,421.9*** |
| Household size 5 (HH5) | 0.05 | -0.01 | -109.6 | 1,975.9*** | 54,215.4*** | 48,938.0*** |
| E | -0.1*** | -0.1*** | 2,184.3*** | 2,420.5*** | 1,838.7 | 955.7 |
| I | 0.1*** | 0.1** | 740.0*** | 1,025.1*** | 743.4 | 307.0 |
| $\tau$ | -0.1 | -0.1 | 2,479.3 | 2,479.3 | 127,568.5*** | 127,568.5*** |
| $\beta$ | 0.5 | 0.5 | -59,516.0*** | -59,516.0*** | -660,496.1*** | -660,496.1*** |
| $\omega$ | -0.7 | -0.7 | -2,710.5 | -2,710.5 | 15,691,158.0*** | 15,691,158.0*** |
| E*HH5 |  | -0.04 |  | -472.6** |  | 1,766.0 |
| I*HH5 |  | 0.1* |  | -570.2*** |  | 872.7 |
| HH3 + HH5 | -0.3 | -0.3 | 25,537.0*** | 25,537.0*** | 190,998.6*** | 190,998.6*** |

Note: \*p<0.1; \*\*p<0.05; \*\*\*p<0.01
HH3 describes how much the outcome differs if the household structure is excluded from an otherwise similar epidemic where the true household size is 3.
HH5 describes the incremental difference in the outcome if the household structure is excluded from an otherwise similar epidemic where the true household size is 5 instead of 3.
For the models, the total effect of exclusion of household structure involves interactions with other terms.
For example, E\*HH5 describes how the incremental difference depends upon the number of exposed compartments when the true household size is 5 instead of 3.
We estimated the magnitude of the linear combination of the relevant coefficients and tested their significance.
E: Number of exposed compartments; I: Number of infectious compartments; $\tau$ : Household transmission rate; $\beta$ : Community transmission rate; $\omega$ : Waning immunity rate.

### 4.3 Impact on model-predicted NPI effects

Failure to include household structure results in substantial biases in model-estimated effects of NPIs, which interact in complex ways with models that have multiple exposed and infectious compartments (Appendix Tables A5 - A6). Panel A of Figure 3 shows how, in the example, the model-estimated reduction in peak size due to NPIs without household structure (Δ*O*_*NH*_) is 32,900 larger than the model-estimated reduction with a household size of 3 (Δ*O*_*HH*_).

**Figure 3:**
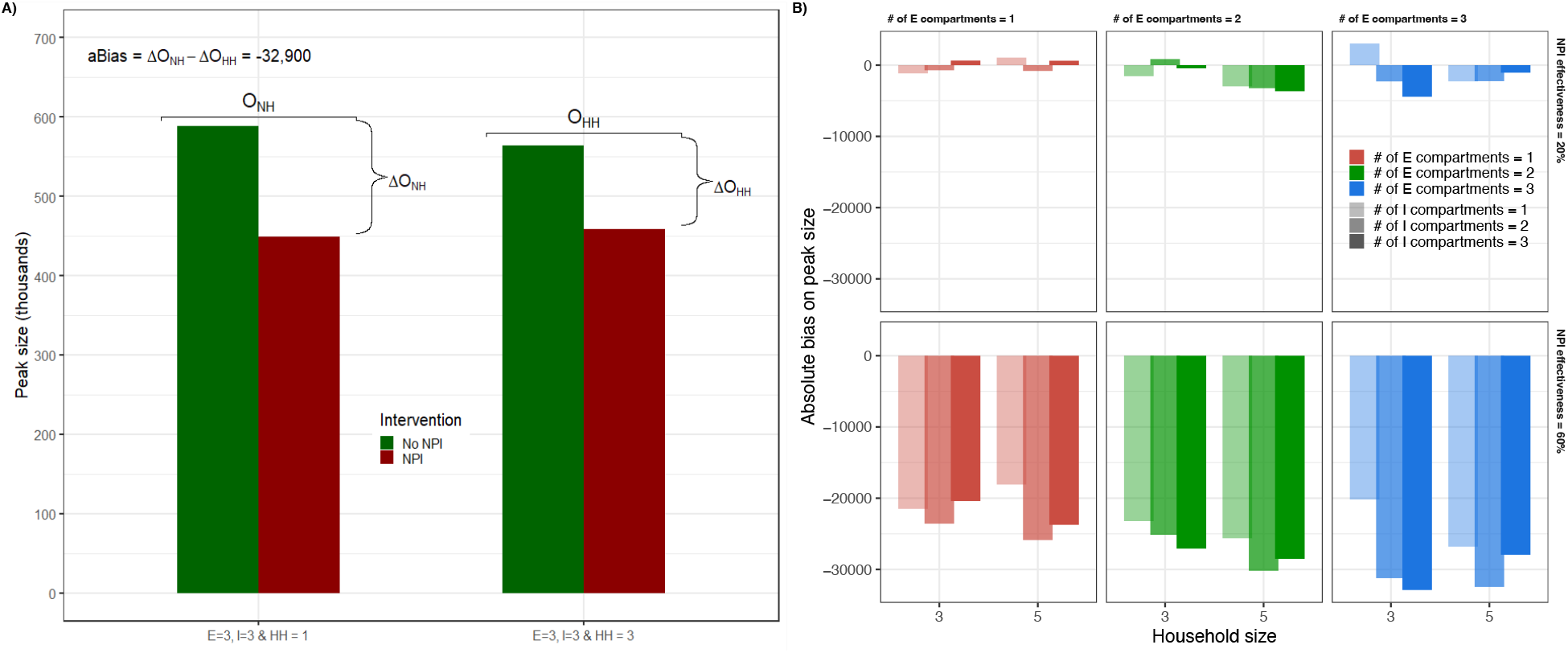
Control measures’ bias. Panel A shows bias with and without household structure for an NPI with 60% effectiveness in reducing community contacts

Using meta-analytic regressions, we found that, on average, model-estimated NPI effects of a 60% reduction in community contacts on peak time and peak size are systematically overestimated when the household structure is not included. In our example, for the model without a household structure, its peak is more delayed by the NPI (approximately 4 days more delayed) than the delay in an otherwise similar model with the household structure. The bias on peak time is slightly smaller when the true average household size is larger. The bias on peak size reduction due to the NPI is larger when the true household size is larger (preventing approximately 44,000 more cases at the peak without household structure when the true household size is 3; preventing approximately 45,000 more cases when the true household size is 5). For overall epidemic size over 100 days, the exclusion of household structure results in an underestimate of the effect of an NPI that reduces community contacts by 60% (approximately 61,000 and 57,000 fewer cases prevented in the first 100 days when the true model’s household size is 3 and 5 respectively) (Table 2). Because models without household structure estimate that NPIs delay peaks longer and push the peak size lower, the effect on the epidemic size over 100 days appears smaller because it will take longer for the epidemic to essentially die out – the key is that the dynamics are different with interventions in the models with and without household structure. Panel B of Figure 3 illustrates this for exemplar parameter sets, showing that the model-estimated effects of NPIs on reducing peak size are frequently overestimated for NPI effectiveness of 60%. However, particular combinations of household size and NEDT exposed and infectious states interact in complex ways to determine the magnitude of this bias.

**Table 2:** Meta-regression estimates on the absolute bias of treatment effects, NPI = 60%.

|  | Peak time (days) |  | Peak size (people) |  | Epidemic size (people) |  |
| --- | --- | --- | --- | --- | --- | --- |
| Household size 3 (HH3) | 4.2*** | 4.3*** | -43,186.4*** | -44,199.8*** | 60,316.3*** | 60,602.9*** |
| Household size 5 (HH5) | -0.4** | -0.6 | -1,825.7*** | 201.2 | -3,513.0*** | -4,086.2*** |
| E | 0.4*** | 0.4*** | -3,428.5*** | -2,600.5*** | 21.5 | 82.4 |
| I | -0.3*** | -0.3** | -1,724.5*** | -2,045.8*** | -1,666.5*** | -1,870.7*** |
| $\tau$ | -8.1*** | -8.1*** | 103,247.7*** | 103,247.7*** | -136,101.6*** | -136,101.6*** |
| $\beta$ | -0.3 | -0.3 | 6,437.3 | 6,437.3 | -4,638.0* | -4,638.0* |
| E*HH5 |  | 0.1 |  | -1,656.1** |  | -121.8 |
| I*HH5 |  | 0.0 |  | 642.6 |  | 408.4 |
| HH3 + HH5 | 3.8*** | 3.8*** | -45,012.1*** | -45,012.1*** | 56,803.3*** | 56,803.3*** |

Note: \*p<0.1; \*\*p<0.05; \*\*\*p<0.01
Absolute bias is equal to effect without HH structure minus effect with structure.
Interventions increase time to peak; Positive bias means the model without HH structure yields a larger effect.
Interventions decrease the size of the peak; Negative bias means the model without HH structure yields a larger effect.
Interventions decrease epidemic size; Negative bias means the model without HH structure yields a larger effect.
HH3 describes how much the intervention effect differs if the household structure is excluded from an otherwise similar epidemic where the true household size is 3.
HH5 describes the incremental difference in the intervention effect if the household structure is excluded from an otherwise similar epidemic where the true household size is 5 instead of 3.
For the models, the total effect of exclusion of household structure involves interactions with other terms.
For example, E\*HH5 describes how the incremental difference depends upon the number of exposed compartments when the true household size is 5 instead of 3.
We estimated the magnitude of the linear combination of the relevant coefficients and tested their significance.

As the NPI less effectively reduces community contacts (e.g., NPI effectiveness of 20%), the biases described above tend to be smaller and non-significant or even to underestimate the reductions in peak size using the exemplar parameter sets (Panel B of Figure 3 and Appendix Figure A.1). Using meta-analytic regressions, we found that, on average, model-estimated NPI effects of a 20% reduction in community contacts on impacting peak time and peak size are no longer significantly biased. For these outcomes as well as the bias in the effect on the total epidemic size, which is still significant, the magnitudes of their point estimates of biases are smaller (tending towards 0) compared to biases for a 60% effective NPI) (Appendix Table A7).

The NPI results presented thus far are for models without waning immunity (*ω* = 0.00). Additional results for models with waning immunity are described in the Appendix.

### 4.4 Impact on model-predicted vaccination effects

Biases from excluding household transmission in the estimates of NPIs’ effects on epidemic outcomes are often larger than the biases in the estimates of the effects of vaccines because the modeled NPIs only impact community transmission. In contrast, vaccination impacts both community and household transmission (Appendix Tables A5 - A6). For a detailed description of the biases from excluding household transmission in the estimates of vaccination, please see the Appendix.

## 5 Discussion

Transmission dynamic models can support policymakers, providing timely epidemic forecasts and assessing the potential effectiveness of multiple control measures. While compartmental transmission dynamic models yield important insights, their simplifying assumptions regarding mixing and transmission can induce biases in epidemic forecasts and estimates of intervention effectiveness, particularly in situations with differential community and household transmission. However, using dynamic transmission microsimulation models may not be feasible because they require too much-unobserved data (e.g., network structure) and computational resources to provide timely results. Our study provides a feasible alternative to address simulation needs within the spectrum of complexity from simple compartmental models to dynamic transmission microsimulation models. Specifically, it extends previous work incorporating household transmission into simple compartmental SIR models [HK08; HK09] to include multi-compartment SEIR models that can include both NPIs that reduce community transmission and vaccination that reduces both community and household transmission. To enable reproducibility and application, we provide an open-source implementation of the modeling framework (https://github.com/SC-COSMO/hhmcseirv).

We demonstrate the value of our framework by comparing simulation results using the framework that incorporates household transmission to simulation results in which household transmission is not explicitly modeled. Across a range of parameters representing diverse pathogens in many epidemiological and social situations, we show that failure to explicitly include household transmission in the model induces bias in its forecasts, particularly on the size of the epidemic peak. Likewise, we show that failure to include household transmission biases estimates of NPI effects on the epidemic outcomes in complex ways. These biases differ substantially and systematically from biases in estimates of vaccine effects. Consistent with empirical studies[Fun+20; Mad+22; Dah+21; Gly+18], the fraction of overall transmission accounted for by within-household transmission varies by parameters representing the setting-specific transmission of the pathogen of interest (i.e., *β* and *τ*) as well as the size of households in the population; likewise, for a fixed set of parameters, this fraction varies over the course of the epidemic.

Failure to include household structure not only induces biases in the modeled overall course of an epidemic and the effects of interventions delivered differentially in community settings. It also limits the ability of the model to evaluate household-specific interventions convincingly. For example, contact investigations could be examined by increasing the rate of detection (and treatment or prophylaxis) among household contacts. Hence, our framework provides additional advantages regarding the types of targeted interventions that can be evaluated.

Our results are consistent with prior literature examining modeled epidemic spread and modeled intervention effects comparing network models to systems of ODEs. These studies have found that network model-predicted timing and size of epidemics differ from those predicted by ODE models and, likewise, that predicted intervention effects in network models would diverge from those of ODE models that produced similar endemic equilibria without intervention. Notably, the magnitude of divergence depends on various parameters, including the degree of clustering in the network models, which may be analogized to the household structure our framework approximates.[SJ10; Mal+21]

Our modeling framework falls within a complexity spectrum between compartmental and microsimulation models. It introduces a set of differential equations whose size grows with the number of modeled states and household size to track households and household transmission. To keep the problem tractable, it assumes that all households are the same size – the average size for the population. Relaxing this assumption would add more equations to the model’s household subcomponent. Solving large systems of ordinary differential equations eventually results in numerical imprecision for compartments with extremely small proportions of the population and requires longer computation times. In our R implementation of the framework, household sizes of more than 5 combined with multi-compartments of more than 4 exposed states and more than 4 infectious states resulted in computation times of minutes to hours. Re-implementing the framework in a high-performance language like C++ or Julia and using appropriate differential equation solvers would likely raise these limits considerably. Even so, dynamic microsimulation models would become preferable at some point of complexity.

As famously noted, “all models are wrong, but some models are useful” [Box76]; the goal of this study is to extend the usefulness of compartmental dynamic transmission models for forecasting and policy evaluation by developing methods to incorporate household transmission into a broad class of such models. We provide the full mathematical details of such an approach and show that incorporating such dynamics into many models is feasible and bias-reducing. We believe that it is advisable to incorporate household transmission into a wide range of dynamic transmission models. With the release of an open-source modeling framework to support analysts, we believe it is advisable to incorporate household transmission into a wide range of dynamic transmission models in the future.

## Data Availability

All data produced in the present study are available upon reasonable request to the authors

https://github.com/SC-COSMO/hhmcseirv

## References

[Box76] George E. P. Box. “Science and Statistics”. In: Journal of the American Statistical Association 71.356 (Dec. 1976), pp. 791–799. ISSN: 0162-1459. DOI: 10.1080/01621459.1976.10480949. URL: http://www.tandfonline.com/doi/abs/10.1080/01621459.1976.10480949.

[HK08] Thomas House and Matt J Keeling. “Deterministic epidemic models with explicit household structure”. In: Mathematical Biosciences 213.1 (2008), pp. 29–39. ISSN: 0025-5564. DOI: 10.1016/j.mbs.2008.01.011. URL: http://www.sciencedirect.com/science/article/pii/S0025556408000199.

[KR08] Matthew James Keeling and Pejman Rohani. Modeling Infectious Diseases in Humans and Animals. Princeton, N.J.: Princeton University Press, 2008, p. 385. ISBN: 9780691116174. URL: http://homepages.warwick.ac.uk/%7B%5C%%7D7B%7B%7D%7B%5C%%7D7Dmasfz/ModelingInfectiousDiseases/index.html.

[Mos+08] Joel Mossong et al. “Social Contacts and Mixing Patterns Relevant to the Spread of Infectious Diseases”. In: PLOS Medicine 5.3 (2008), p. 1. DOI: 10.1371/journal.pmed.0050074. URL: https://doi.org/10.1371/journal.pmed.0050074.

[HK09] T House and M J Keeling. “Household structure and infectious disease transmission.” eng. In: Epidemiology and infection 137.5 (May 2009), pp. 654–661. ISSN: 0950-2688 (Print). DOI: 10.1017/S0950268808001416.

[SJ10] Marcel Salathe and James H. Jones. “Dynamics and Control of Diseases in Networks with Community Structure”. In: PLoS Computational Biology 6.4 (Apr. 2010). Ed. by Christophe Fraser, e1000736. ISSN: 1553-7358. DOI: 10.1371/journal. pcbi. 1000736. URL: https://dx.plos.org/10.1371/journal.pcbi.1000736.

[HKP17] Michael Harwell, Nidhi Kohli, and Yadira Peralta. “Experimental Design and Data Analysis in Computer Simulation Studies in the Behavioral Sciences”. In: Journal of Modern Applied Statistical Methods 16.2 (2017), pp. 3–28. ISSN: 1538-9472. DOI: 10.22237/jmasm/1509494520. URL: http://digitalcommons.wayne.edu/jmasm/vol16/iss2/2 http://digitalcommons.wayne.edu/cgi/viewcontent.cgi?article=2531&context=jmasm.

[Gly+18] Judith R. Glynn et al. “Variability in Intrahousehold Transmission of Ebola Virus, and Estimation of the Household Secondary Attack Rate”. In: Journal of Infectious Diseases 217.2 (2018), pp. 232–237. ISSN: 15376613. DOI: 10.1093/infdis/jix579.

[MWC19] Tim P Morris, Ian R White, and Michael J Crowther. “Using simulation studies to evaluate statistical methods”. In: Statistics in Medicine 38.11 (2019), pp. 2074–2102. ISSN: 02776715. DOI: 10.1002/sim.8086. arXiv: 1712.03198. URL: http://arxiv.org/abs/1712.03198.

[Fun+20] Hannah F Fung et al. “The household secondary attack rate of SARS-CoV-2: A rapid review”. In: Clinical Infectious Diseases (2020). URL: https://academic.oup.com/cid/advance-article/doi/10.1093/cid/ciaa1558/5921151.

[Pel+20] Lorenzo Pellis et al. “Systematic selection between age and household structure for models aimed at emerging epidemic predictions”. In: Nature Communications 11.1 (2020).

[Ala+21] Fernando Alarid-Escudero et al. “Dependence of COVID-19 Policies on End-of-Year Holiday Contacts in Mexico City Metropolitan Area: A Modeling Study”. In: Medical Decision Making Policy & Practice 6.2 (2021), pp. 1–14. DOI: 10.1177/23814683211049249. URL: https://journals.sagepub.com/doi/full/10.1177/23814683211049249.

[Dah+21] F. Scott Dahlgren et al. “Household transmission of influenza A and B within a prospective cohort during the 2013-2014 and 2014-2015 seasons”. In: Statistics in Medicine 40.28 (2021), pp. 6260–6276. ISSN: 10970258. DOI: 10.1002/sim.9181.

[Mal+21] Giovanni S.P. Malloy et al. “Predicting the Effectiveness of Endemic Infectious Disease Control Interventions: The Impact of Mass Action versus Network Model Structure”. In: Medical Decision Making 41.6 (2021), pp. 623–640. ISSN: 1552681X. DOI: 10.1177/0272989X211006025.

[Dat22] Harvard Dataverse. World COVID-19 Daily Cases with Basemap. en. June 2022. DOI: 10.7910/DVN/L20LOT. URL: https://dataverse.harvard.edu/dataset.xhtml?persistentId=doi:10.7910/DVN/L20LOT (visited on 05/18/2023).

[Mad+22] Zachary J. Madewell et al. “Household Secondary Attack Rates of SARS-CoV-2 by Variant and Vaccination Status: An Updated Systematic Review and Meta-analysis”. In: JAMA Network Open 5.4 (2022), E229317. ISSN: 25743805. DOI: 10.1001/jamanetworkopen.2022.9317.

[Dat23] Harvard Dataverse. US COVID-19 Daily Cases with Basemap. en. May 2023. DOI: 10.7910/DVN/HIDLTK. URL: https://dataverse.harvard.edu/dataset.xhtml?persistentId=doi:10.7910/DVN/HIDLTK (visited on 05/18/2023).

[Goo23] Google. COVID-19 Community Mobility Report. May 2023. URL: https://www.google. com/covid19/mobility?hl=en (visited on 05/18/2023).

