## Appendix for "Effects of Mitigation and Control Policies in Realistic Epidemic Models Accounting for Household Transmission Dynamics"

### A.1 Technical details

#### A.1.1 Community model equation

The community MC-SEIRSV model is described by a system of $[3+J+2K]$ ordinary differential equations (ODEs):

$$\begin{aligned}
\frac{dS}{dt} &= bN + \omega R + \omega_v V - \left(\phi\lambda(t) + \lambda_{HH}(t) + \mu + h\theta\right) S \\
\frac{dE^1}{dt} &= \left(\phi\lambda(t) + \lambda_{HH}(t)\right) S - \left(\sigma J + \mu\right) E^1 \\
\frac{dE^j}{dt} &= \sigma J E^{j-1} - \left(\sigma J + \mu\right) E^j, \text{ for } j = 2, \ldots, J \\
\frac{dI^1}{dt} &= \sigma J E^J - \left(\gamma K + \nu + \mu\right) I^1 \\
\frac{dI^k}{dt} &= \gamma K I^{k-1} - \left(\gamma K + \nu + \mu\right) I^k, \text{ for } k = 2, \ldots, K \\
\frac{dIDX^1}{dt} &= \nu I^1 - \left(\gamma K + \mu\right) IDX^1 \\
\frac{dIDX^k}{dt} &= \nu I^k + (\gamma K) IDX^{k-1} - \left(\gamma K + \mu\right) IDX^k, \text{ for } k = 2, \ldots, K \\
\frac{dR}{dt} &= \gamma K \left(1 - p^d\right)\left(I^K + IDX^K\right) - (\omega + \mu) R \\
\frac{dV}{dt} &= h\theta S - (\omega_v + \mu) V,
\end{aligned} \tag{5}$$

where $N = S + \sum_{j=1}^{J} E^j + \sum_{k=1}^{K}\left(I^k + IDX^k\right) + R + V$ is the total population; $b$ is the birth rate; $\sigma$ is the rate at which exposed individuals in class $E^j$ progress to class $E^{j+1}$ for $j = 1, \ldots, (J-1)$ and from the exposed class $E^J$ to the infected class $I^1$; $\gamma$ is the rate at which infectious individuals in class $I^k$ progress to class $I^{k+1}$ and also from $IDX^k$ progress to class $IDX^{k+1}$ for $k = 1, \ldots, (K-1)$ and it is also the recovery rate from the infectious classes $I^K$ and $IDX^K$ to the recovered class $R$; $\nu$ is the detection rate from which $I^k$ go to $IDX^k$ for $k = 1, \ldots, K$; $p^d$ is the proportion of infectious individuals in classes $I^K$ and $IDX^K$ that die from the disease; $\omega$ is the rate of waning immunity at which $R$ goes to $S$; and $\mu$ represents background mortality experienced from all compartments. The model accounts for two types of forces of infection, from the community, $\lambda(t)$, and the household, $\lambda_{HH}(t)$, which is determined by the household submodel and, hence, links the household submodel to the community model and keeps them consistent (see 2.2.1 in the main manuscript as well as the pseudo-code of this linkage in A.1.3 in the appendix). We modeled NPIs as a reduction in the community FOI, $\lambda(t)$ by an amount $\phi \in [0, 1]$. Susceptible individuals get vaccinated at a rate $h$ for whom a proportion, $\theta \in [0, 1]$, the vaccines will be effective. Vaccinated individuals will face their immunity waning at a rate $\omega_v$.

#### A.1.2 Community force of infection

The community FOI is defined as

$$\lambda(t) = \beta \sum_{k=1}^{K} \frac{I^k(t) + fIDX^k(t)}{N}, \tag{6}$$

where the transmission rate, $\beta$, describes the probability that an infected individual who is $k$ days into the infectious period will infect a susceptible per unit of time, and $f \in [0, 1]$ is a reduction factor in transmission from infectious individuals that are diagnosed (due for example to quarantine and isolation).

##### A.1.3 Pseudo-code for linkage between the Community and Households submodels

For each time $t$ from 0 to $T$:

1. Community submodel requests household force of infection from the household submodel (Lines 204-212 in file model_functions.R)
   (a) Household submodel computes household force of infection as in equation 2 in the main text. The code generates the weighted averaged of the force of infection for households in all possible state counts (numbers of members in each given MC-SEIRV state) where the weights are the proportions of households in the state counts and the number of susceptible individuals in them. (Lines 358-367 in model_functions.R)
2. Community submodel computes community force of infection directly. (Line 217 in model_functions.R)
3. Community submodel applies forces of infection from steps 1 and 2 to infect the appropriate proportion of the overall population in S for time $t + \Delta t$. (Lines 226-227 in model_functions.R)
4. Household submodel applies the community force of infection to each susceptible individual in households with one or more susceptible individuals to compute the new proportions of households with different state counts for time $t + \Delta t$ as described in A.1.5 (Lines 271-272 in model_functions.R)

While the actual models are solved in continuous time, for simplicity, the pseudo-code above describes the algorithm in discrete time steps, though $\Delta t$ can be arbitrarily small.

##### A.1.4 Births and Deaths

Like the community submodel, the household submodel includes birth and deaths. Because the household submodel tracks proportions, it makes the simplifying assumption that births equal deaths so that its proportions always sum to 1, $\sum_{sc} HH_{sc} = 1$, and that at any given time, the fraction of incident deaths due to disease is relatively small. Hence, it exposes all households to an average background mortality rate, determining the birth rate. Births are spread proportionally across only household compartments with at least one susceptible member (i.e., newly born individuals are assumed to be born susceptible). This assumption could be relaxed if vertical transmission occurred.

##### A.1.5 Formal construction of the distribution of household members in source and destination states at times $t$ and $t + 1$

Consider $C$ individuals (i.e., the members of a household) each in a Markov chain with states $X_t^c \in \{1, \ldots, M\}$ for $c \in \{1, \ldots, C\}$. The $M$ states in our case are those in the community MC-SEIRSV model. The Markov chain has the following transition probabilities (where for simplicity, we set the probability of flow from $R$ to $S$ equal to 0, i.e., no waning immunity):

$$P(X^c_{t+1} = X^c_t + 1 \mid X^c_t) = p, \quad X_t = 1, \ldots, M-1 \tag{7}$$

$$P(X^c_{t+1} = X^c_t \mid X^c_t) = 1 - p, \quad X_t = 1, \ldots, M-1 \tag{8}$$

$$P(X^c_{t+1} = X^c_t \mid X^c_t) = 1, \quad X_t = M \tag{9}$$

In other words, for all states except the last, with probability $p$, each individual progresses from $X_t$ to $X_{t+1}$ (e.g., $E_2$ to $E_3$ or from $E_3$ to $I_1$) and with probability $1-p$ the individual stays in the same state. Individuals remain with certainty in the last ($M^{th}$) state after progressing to it.

Having considered each individual, we now consider counts of household members. Let $Y^m_t$ be the number of individuals in state $m$ at time $t$. We consider a new Markov chain with state $(Y^1_t, \ldots, Y^M_t)$. The transition probabilities for the counts of household members can be calculated as follows. Given a transition from state $(Y^1_t, \ldots, Y^M_t)$ to state $(Y^1_{t+1}, \ldots, Y^M_{t+1})$:

1. For each of the $M-1$ transition arcs in the underlying Markov chain, find the number of individuals transitioning from state $m$ to $m+1$, denoted $\Delta_t(m, m+1)$ for $m < M$.

2. The probability of the transition is then a (convolution of) binomial distribution(s):

$$\prod_{m=1}^{M-1} \binom{Y^m_t}{\Delta_t(m, m+1)} p^{\Delta_t(m,m+1)} (1-p)^{Y^m_t - \Delta_t(m,m+1)} \tag{10}$$

To find the value of $\Delta_t(m, m+1)$, use the following backwards recursion:

$$\Delta_t(M-1, M) = Y^M_{t+1} - Y^M_t \tag{11}$$

$$\Delta_t(m-1, m) = Y^m_{t+1} + \Delta_t(m, m+1) - Y^m_t \tag{12}$$

##### A.1.6 Definition of epidemic outcomes

We define multiple epidemic outcomes, $O$, that we analyze in this study. These include the cumulative epidemic size, $ES$; the size of the epidemic peak, $EP$; and the timing of the epidemic peak, $tEP$.

Prevalent infections at time $t$, $PI$, are defined as the sum of all exposed, $E^j$ for $j = 1, \ldots, J$, and undiagnosed and diagnosed infectious individuals, $I^k$ and $IDX^k$ for $k = 1, \ldots, K$, respectively, given by

$$PI = \sum_{j=1}^{J} E^j + \sum_{k=1}^{K} \left( I^k + IDX^k \right). \tag{13}$$

The cumulative infection time up to a time $T$, representing the epidemic size, $ES$, is defined as

$$ES = \int_{t=0}^{T} PI(t)dt. \tag{14}$$

We quantified the magnitude of the epidemic as the epidemic peak, $EP$, defined as the highest number of prevalent infections over a specified period $T$, given by

$$EP = \max\{PI\} \text{ for } t \in [0, T]. \tag{15}$$

The timing of the epidemic peak, $tEP$, is defined as the time $t$ at which the epidemic peak occurs over a specified period $T$, given by

$$tEP = \arg\max_t \{PI\} \text{ for } t \in [0, T]. \tag{16}$$

### A.2 Model parameters

Table A1: Model parameters

| Parameter | Base-case value | DOE values |
|---|---|---|
| Number of exposed compartments (E) | 3 | 1-3 |
| Number of infectious compartments (I) | 2 | 1-3 |
| Household size (HH) | 3 | 1-5 |
| Household transmission rate ($\tau$) | 0.40 | 0.40, 0.50 |
| Community transmission rate ($\beta$) | 0.25 | 0.25, 0.35 |
| Waning immunity rate ($\omega$) | 0.00 | 0.01, 0.02 |
| Progression rate ($\sigma$) | 0.35 | - |
| Recovery rate ($\gamma$) | 0.20 | - |
| Excess risk of death from infection $\left(p^d\right)$ | 0.02 | - |
| Reduced transmission in diagnosed I ($f$) | 0.20 | - |
| NPI effectiveness ($1 - \phi$) | 0.0 | 0.0, 0.2, 0.6 |
| Vaccine effectiveness ($\theta$) | 1.0 | 1.0, 0.9, 0.5 |
| Vaccination coverage (%) | 0 | 0, 20, 60, 90 |

*Notes:*
DOE: Design of experiment

#### A.3 Calibration results

Table A2: Metaregression estimates of how much larger $\beta$ is for otherwise similar models without household structure

| Parameter | Effect |
|---|---|
| $\tau$ | 0.223*** |
| $\beta$ | 0.278*** |
| $\omega$ | 0.007 |
| E | 0.002*** |
| I | -0.001*** |
| Household size 3 (HH3) | -0.028*** |
| Household size 5 (HH5) | 0.126*** |
| *Note:* | *p<0.1; **p<0.05; ***p<0.01 |

#### A.4 Additional results

#### A.5 Impact on model-predicted NPI effects with waning immunity

Considering analyses of intervention effect bias for ($\omega = 0.01$ or $\omega = 0.02$) for 60% NPI effectiveness in reducing community contacts, we find that for the case of $\omega = 0.01$ biases on peak time and peak size are only slightly larger than for the case of $\omega = 0.00$. For overall epidemic size within 100 days, the bias appears much larger (approximately 600,000 vs. 60,000 total fewer cases averted for models without household structure) when $\omega = 0.01$ compared to $\omega = 0.00$. This is a combination of many more cases occurring because waning immunity permits those with prior infection to become susceptible again and changes in the timing of both the epidemic peak and when the endemic equilibrium is reached (Appendix Table A8). For $\omega = 0.02$, biases on peak time and peak size are slightly smaller than for the case of $\omega = 0.00$ but generally similar and still in the same direction. However, for total epidemic size, the direction of bias has now changed (approximately -550,000 vs. 60,000 total fewer cases averted for models without household structure) – with $\omega = 0.02$, the model without household structure overestimates the reduction in total cases within 100 days (Appendix Table A9).

#### A.6 Impact on model-predicted vaccination effects

Biases from excluding household transmission in the estimates of NPIs' effects on epidemic outcomes are often larger than the biases in the estimates of the effects of vaccines because the modeled NPIs only impact community transmission. In contrast, vaccination impacts both community and household transmission (Appendix Tables A5 - A6). Appendix Figures A.2 and A.3 show examples of how vaccines with given effectiveness and coverage produce different epidemic curves with and without including household transmission holding all other model parameters fixed. For this example, the effect on peak size and timing is smaller for models with realistic household sizes and leads to less bias than in estimates with NPI effects (Appendix Figure A.5 compared to Figure 3). More generally, we see that estimated effects on our outcomes of low coverage and low effectiveness vaccine from models without household structure compared to otherwise similar models with household structure yields no significant bias on average (Appendix Table A10). With a low coverage but high effectiveness vaccine, the findings are similar, though, for peak size, the model without household structure may underestimate the effect of the vaccine by approximately 9,000 compared to otherwise similar models with household structure (Appendix Table A11). However, for high-effective vaccine strategies at low or high coverage, different patterns of bias emerge due to models with and without household structure having different thresholds for elimination (Appendix Tables A12 and A13). Estimated delays on peak times are significantly smaller for models without household structure compared to those with household structure when vaccine coverage is lower but no longer significant with higher vaccine coverage. For peak time, models without household structure significantly underestimate reductions with lower vaccine coverage (approximately 16,000) but significantly overestimate reductions with higher coverage (approximately 17,000). For low and high coverage, reductions in total epidemic size are overestimated in models without household structure compared to those with household structure.

Table A3: Outcomes for models with and without household structure, household size = 3

| Scenario | HH | Peak Time (SD) | Peak Infections (SD) | Epidemic Size (SD) |
|---|---|---|---|---|
| Nat Hx | FALSE | 20 (3) | 524,200 (52,500) | 5,000,000 (0) |
| Nat Hx | TRUE | 20 (3) | 509,400 (51,600) | 4,999,900 (100) |
| NPI=60% | FALSE | 25 (4) | 426,000 (46,500) | 4,994,500 (4,300) |
| NPI=60% | TRUE | 23 (4) | 430,800 (46,300) | 4,980,300 (12,200) |
| NPI=20% | FALSE | 21 (3) | 507,800 (51,000) | 5,000,000 (0) |
| NPI=20% | TRUE | 21 (3) | 493,500 (50,500) | 4,999,500 (500) |
| Vax Prop=30%; Eff=50% | FALSE | 20 (3) | 431,000 (43,700) | 4,209,100 (40,600) |
| Vax Prop=30%; Eff=50% | TRUE | 21 (3) | 413,200 (43,200) | 4,210,800 (44,500) |
| Vax Prop=90%; Eff=50% | FALSE | 24 (5) | 134,000 (28,100) | 1,670,900 (152,700) |
| Vax Prop=90%; Eff=50% | TRUE | 26 (6) | 111,200 (32,500) | 1,641,000 (176,600) |
| Vax Prop=30%; Eff=90% | FALSE | 21 (3) | 367,300 (38,100) | 3,669,600 (66,900) |
| Vax Prop=30%; Eff=90% | TRUE | 21 (3) | 347,800 (38,500) | 3,671,700 (73,100) |
| Vax Prop=90%; Eff=90% | FALSE | 29 (9) | 35,900 (27,700) | 664,800 (206,700) |
| Vax Prop=90%; Eff=90% | TRUE | 32 (14) | 24,900 (30,000) | 525,100 (287,400) |
| Nat Hx; Omega=0.01 | FALSE | 20 (3) | 525,300 (52,300) | 7,787,700 (142,500) |
| Nat Hx; Omega=0.01 | TRUE | 20 (3) | 510,500 (51,300) | 7,586,000 (160,600) |
| NPI=60%; Omega=0.01 | FALSE | 25 (4) | 428,100 (46,000) | 6,973,400 (367,700) |
| NPI=60%; Omega=0.01 | TRUE | 23 (4) | 432,700 (45,900) | 6,624,300 (502,900) |
| Nat Hx; Omega=0.02 | FALSE | 20 (3) | 526,300 (52,000) | 10,437,800 (237,400) |
| Nat Hx; Omega=0.02 | TRUE | 20 (3) | 511,600 (51,000) | 10,165,200 (282,300) |
| NPI=60%; Omega=0.02 | FALSE | 25 (4) | 430,200 (45,600) | 9,303,000 (392,600) |
| NPI=60%; Omega=0.02 | TRUE | 24 (4) | 434,500 (45,500) | 9,098,500 (365,100) |

Table A4: Outcomes for models with and without household structure, household size = 5

| Scenario | HH | Peak Time (SD) | Peak Infections (SD) | Epidemic Size (SD) |
|---|---|---|---|---|
| Nat Hx | FALSE | 18 (2) | 538,500 (54,300) | 5,000,000 (0) |
| Nat Hx | TRUE | 18 (2) | 523,900 (53,700) | 4,999,900 (100) |
| NPI=60% | FALSE | 21 (4) | 463,400 (48,300) | 4,999,100 (800) |
| NPI=60% | TRUE | 20 (3) | 470,200 (48,800) | 4,988,500 (7,100) |
| NPI=20% | FALSE | 18 (3) | 526,000 (53,100) | 5,000,000 (0) |
| NPI=20% | TRUE | 18 (3) | 512,500 (52,900) | 4,999,700 (300) |
| Vax Prop=30%; Eff=50% | FALSE | 18 (3) | 453,500 (46,800) | 4,278,800 (144,100) |
| Vax Prop=30%; Eff=50% | TRUE | 18 (3) | 434,700 (47,100) | 4,282,800 (146,300) |
| Vax Prop=90%; Eff=50% | FALSE | 20 (4) | 180,000 (71,600) | 1,943,600 (555,700) |
| Vax Prop=90%; Eff=50% | TRUE | 21 (5) | 152,200 (79,300) | 1,922,700 (568,900) |
| Vax Prop=30%; Eff=90% | FALSE | 18 (3) | 395,300 (46,500) | 3,785,400 (239,200) |
| Vax Prop=30%; Eff=90% | TRUE | 19 (3) | 373,800 (48,400) | 3,790,700 (242,200) |
| Vax Prop=90%; Eff=90% | FALSE | 21 (6) | 83,600 (84,100) | 1,032,200 (673,400) |
| Vax Prop=90%; Eff=90% | TRUE | 22 (8) | 67,900 (88,400) | 917,900 (738,700) |

| | | | | |
|---|---|---|---|---|
| Nat Hx; Omega=0.01 | FALSE | 18 (2) | 539,400 (54,000) | 7,966,200 (119,900) |
| Nat Hx; Omega=0.01 | TRUE | 18 (2) | 524,800 (53,400) | 7,684,500 (152,000) |
| NPI=60%; Omega=0.01 | FALSE | 21 (4) | 465,000 (47,900) | 7,397,400 (191,100) |
| NPI=60%; Omega=0.01 | TRUE | 20 (3) | 471,600 (48,400) | 7,090,800 (296,600) |
| Nat Hx; Omega=0.02 | FALSE | 18 (2) | 540,300 (53,800) | 10,711,400 (204,800) |
| Nat Hx; Omega=0.02 | TRUE | 18 (2) | 525,700 (53,200) | 10,356,200 (249,300) |
| NPI=60%; Omega=0.02 | FALSE | 21 (4) | 466,700 (47,400) | 9,843,200 (341,400) |
| NPI=60%; Omega=0.02 | TRUE | 20 (3) | 473,000 (48,000) | 9,478,800 (320,900) |

Table A5: Impact on outcomes relative to no intervention for models with and without household structure, household size = 3

| Scenario | HH | Diff. Peak Time (SD) | Diff. Peak Infections (SD) | Diff. Epidemic Size (SD) |
|---|---|---|---|---|
| NPI=60% | FALSE | 5 (2) | 426,000 (46,500) | -5,500 (4,300) |
| NPI=60% | TRUE | 3 (1) | 430,800 (46,300) | -19,600 (12,100) |
| NPI=20% | FALSE | 1 (0) | 507,800 (51,000) | 0 (0) |
| NPI=20% | TRUE | 1 (0) | 493,500 (50,500) | -400 (400) |
| Vax Prop=30%; Eff=50% | FALSE | 0 (0) | 431,000 (43,700) | -790,900 (40,600) |
| Vax Prop=30%; Eff=50% | TRUE | 1 (1) | 413,200 (43,200) | -789,100 (44,400) |
| Vax Prop=90%; Eff=50% | FALSE | 4 (2) | 134,000 (28,100) | -3,329,100 (152,700) |
| Vax Prop=90%; Eff=50% | TRUE | 6 (4) | 111,200 (32,500) | -3,358,900 (176,600) |
| Vax Prop=30%; Eff=90% | FALSE | 1 (1) | 367,300 (38,100) | -1,330,400 (66,900) |
| Vax Prop=30%; Eff=90% | TRUE | 1 (1) | 347,800 (38,500) | -1,328,200 (73,000) |
| Vax Prop=90%; Eff=90% | FALSE | 9 (7) | 35,900 (27,700) | -4,335,200 (206,700) |
| Vax Prop=90%; Eff=90% | TRUE | 12 (11) | 24,900 (30,000) | -4,474,800 (287,400) |
| NPI=60%; Omega=0.01 | FALSE | 5 (2) | 428,100 (46,000) | -814,300 (230,600) |
| NPI=60%; Omega=0.01 | TRUE | 3 (1) | 432,700 (45,900) | -961,700 (346,700) |
| NPI=60%; Omega=0.02 | FALSE | 5 (2) | 430,200 (45,600) | -1,134,700 (161,800) |
| NPI=60%; Omega=0.02 | TRUE | 3 (1) | 434,500 (45,500) | -1,066,700 (85,000) |

Table A6: Impact on outcomes relative to no intervention for models with and without household structure, household size = 5

| Scenario | HH | Diff. Peak Time (SD) | Diff. Peak Infections (SD) | Diff. Epidemic Size (SD) |
|---|---|---|---|---|
| NPI=60% | FALSE | 3 (1) | 463,400 (48,300) | -900 (800) |
| NPI=60% | TRUE | 2 (1) | 470,200 (48,800) | -11,500 (7,000) |
| NPI=20% | FALSE | 1 (0) | 526,000 (53,100) | 0 (0) |
| NPI=20% | TRUE | 0 (0) | 512,500 (52,900) | -300 (300) |
| Vax Prop=30%; Eff=50% | FALSE | 0 (0) | 453,500 (46,800) | -721,200 (144,100) |
| Vax Prop=30%; Eff=50% | TRUE | 0 (0) | 434,700 (47,100) | -717,100 (146,300) |
| Vax Prop=90%; Eff=50% | FALSE | 2 (2) | 180,000 (71,600) | -3,056,400 (555,700) |
| Vax Prop=90%; Eff=50% | TRUE | 3 (3) | 152,200 (79,300) | -3,077,300 (568,900) |
| Vax Prop=30%; Eff=90% | FALSE | 0 (0) | 395,300 (46,500) | -1,214,600 (239,200) |
| Vax Prop=30%; Eff=90% | TRUE | 1 (1) | 373,800 (48,400) | -1,209,300 (242,100) |
| Vax Prop=90%; Eff=90% | FALSE | 3 (4) | 83,600 (84,100) | -3,967,800 (673,400) |
| Vax Prop=90%; Eff=90% | TRUE | 4 (5) | 67,900 (88,400) | -4,082,000 (738,700) |
| NPI=60%; Omega=0.01 | FALSE | 3 (1) | 465,000 (47,900) | -568,800 (75,800) |
| NPI=60%; Omega=0.01 | TRUE | 2 (1) | 471,600 (48,400) | -593,700 (165,200) |
| NPI=60%; Omega=0.02 | FALSE | 3 (1) | 466,700 (47,400) | -868,200 (143,200) |
| NPI=60%; Omega=0.02 | TRUE | 2 (1) | 473,000 (48,000) | -877,500 (76,200) |

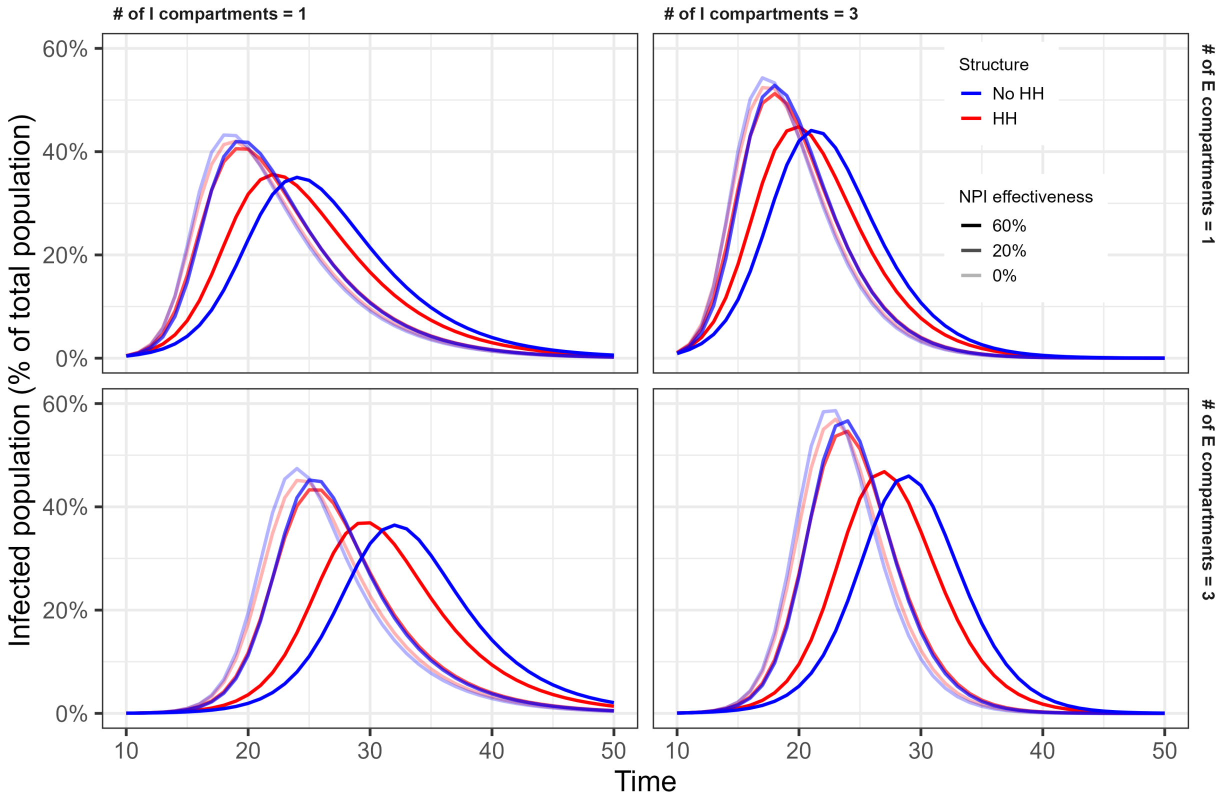


Figure A.1: Epidemic curves under no NPI and NPI effectiveness of 20 or 60%.

Table A7: Meta-regression estimates on absolute bias of treatment effects, NPI = 20%

| | Peak time (days) | | Peak size (people) | | Epidemic size (people) | |
|---|---|---|---|---|---|---|
| Household size 3 (HH3) | 0.4 | 0.5 | -2,185.7 | -2,482.7 | 2,644.0*** | 2,678.3*** |
| Household size 5 (HH5) | 0.1 | -0.1 | -631.9 | -37.9 | -160.3*** | -229.0** |
| E | 0.1 | -0.0 | -563.0* | -327.4 | -7.2 | -2.0 |
| I | -0.02 | -0.0 | -131.4 | -218.5 | -101.0*** | -123.4*** |
| $\tau$ | -1.1 | -1.1 | 8,855.8* | 8,855.8* | -6,217.4*** | -6,217.4*** |
| $\beta$ | -0.0 | -0.0 | 1,019.3 | 1,019.3 | -285.6 | -285.6 |
| E*HH5 | | 0.2 | | -471.3 | | -10.3 |
| I*HH5 | | -0.04 | | 174.4 | | 44.7 |
| HH3 + HH5 | 0.5 | 0.5 | -2,817.6 | -2,817.6 | 2,483.7*** | 2,483.7*** |

*Note:* *p<0.1; **p<0.05; ***p<0.01
Absolute bias is equal to effect without HH structure minus effect with structure.
Interventions increase time to peak; Positive bias means the model without HH structure yields a larger effect.
Interventions decrease the size of the peak; Negative bias means the model without HH structure yields a larger effect.
Interventions decrease epidemic size; Negative bias means the model without HH structure yields a larger effect.
HH3 describes how much the intervention effect differs if the household structure is excluded from an otherwise similar epidemic where the true household size is 3.
HH5 describes the incremental difference in the intervention effect if the household structure is excluded from an otherwise similar epidemic where the true household size is 5 instead of 3.
For the models, the total effect of exclusion of household structure involves interactions with other terms.
For example, E*HH5 describes how the incremental difference depends upon the number of exposed compartments when the true household size is 5 instead of 3.
We estimated the magnitude of the linear combination of the relevant coefficients and tested their significance.
E: Number of exposed compartments; I: Number of infectious compartments; $\tau$: Household transmission rate; $\beta$: Community transmission rate; $\omega$: Waning immunity rate.

Table A8: Meta-regression estimates on absolute bias of treatment effects, NPI = 60% and $\omega$ = 0.01

| | Peak time (days) | | Peak size (people) | | Epidemic size (people) | |
|---|---|---|---|---|---|---|
| Household size 3 (HH3) | 4.2*** | 4.3*** | -42,478.5*** | -43,531.4*** | 658,194.9*** | 588,559.7*** |
| Household size 5 (HH5) | -0.4** | -0.6 | -1,926.2*** | 179.5 | -122,555.2*** | 16,715.3 |
| E | 0.4*** | 0.4*** | -3,403.3*** | -2,568.4*** | 20,279.6* | 30,188.7** |
| I | -0.3*** | -0.3** | -1,824.7*** | -2,133.3*** | -59,693.0*** | -34,784.5** |
| $\tau$ | -8.1*** | -8.1*** | 102,188.5*** | 102,188.5*** | -1,512,210.0*** | -1,512,210.0*** |
| $\beta$ | -0.3 | -0.3 | 6,753.8 | 6,753.8 | 48,225.9 | 48,225.9 |
| E*HH5 | | 0.04 | | -1,669.9** | | -19,818.2 |
| I*HH5 | | 0.04 | | 617.1 | | -49,817.1** |
| HH3 + HH5 | 3.8*** | 3.8*** | -44,404.7*** | -44,404.7*** | 535,639.8*** | 535,639.8*** |

*Note:* *p<0.1; **p<0.05; ***p<0.01
Absolute bias is equal to effect without HH structure minus effect with structure.
Interventions increase time to peak; Positive bias means the model without HH structure yields a larger effect.
Interventions decrease the size of the peak; Negative bias means the model without HH structure yields a larger effect.
Interventions decrease epidemic size; Negative bias means the model without HH structure yields a larger effect.
HH3 describes how much the intervention effect differs if the household structure is excluded from an otherwise similar epidemic where the true household size is 3.
HH5 describes the incremental difference in the intervention effect if the household structure is excluded from an otherwise similar epidemic where the true household size is 5 instead of 3.
For the models, the total effect of exclusion of household structure involves interactions with other terms.
For example, E*HH5 describes how the incremental difference depends upon the number of exposed compartments when the true household size is 5 instead of 3.
We estimated the magnitude of the linear combination of the relevant coefficients and tested their significance.
E: Number of exposed compartments; I: Number of infectious compartments; $\tau$: Household transmission rate; $\beta$: Community transmission rate; $\omega$: Waning immunity rate.

Table A9: Meta-regression estimates on absolute bias of treatment effects, NPI = 60% and $\omega$ = 0.02

| | Peak time (days) | | Peak size (people) | | Epidemic size (people) | |
|---|---|---|---|---|---|---|
| Household size 3 (HH3) | 3.7*** | 3.7*** | -42,047.4*** | -43,168.8*** | -560,427.0*** | -549,945.9*** |
| Household size 5 (HH5) | -0.3* | -0.3 | -1,977.4*** | 265.4 | 77,323.8*** | 56,361.6*** |
| E | 0.4*** | 0.4*** | -3,389.1*** | -2,554.7*** | -32,705.6*** | -32,461.1*** |
| I | -0.3** | -0.2 | -1,894.7*** | -2,168.4*** | 26,811.6*** | 21,326.5*** |
| $\tau$ | -6.9*** | -6.9*** | 101,761.5*** | 101,761.5*** | 1,254,683.0*** | 1,254,683.0*** |
| $\beta$ | -0.3 | -0.3 | 7,026.0 | 7,026.0 | 283,854.4*** | 283,854.4*** |
| E*HH5 | | 0.1 | | -1,668.8** | | -489.1 |
| I*HH5 | | -0.1 | | 547.3 | | 10,970.2** |
| HH3 + HH5 | 3.4*** | 3.4*** | -44,024.7*** | -44,024.7*** | -483,103.2*** | -483,103.2*** |

*Note:* *p<0.1; **p<0.05; ***p<0.01
Absolute bias is equal to effect without HH structure minus effect with structure.
Interventions increase time to peak; Positive bias means the model without HH structure yields a larger effect.
Interventions decrease the size of the peak; Negative bias means the model without HH structure yields a larger effect.
Interventions decrease epidemic size; Negative bias means the model without HH structure yields a larger effect.
HH3 describes how much the intervention effect differs if the household structure is excluded from an otherwise similar epidemic where the true household size is 3.
HH5 describes the incremental difference in the intervention effect if the household structure is excluded from an otherwise similar epidemic where the true household size is 5 instead of 3.
For the models, the total effect of exclusion of household structure involves interactions with other terms.
For example, E*HH5 describes how the incremental difference depends upon the number of exposed compartments when the true household size is 5 instead of 3.
We estimated the magnitude of the linear combination of the relevant coefficients and tested their significance.
E: Number of exposed compartments; I: Number of infectious compartments; $\tau$: Household transmission rate; $\beta$: Community transmission rate; $\omega$: Waning immunity rate.

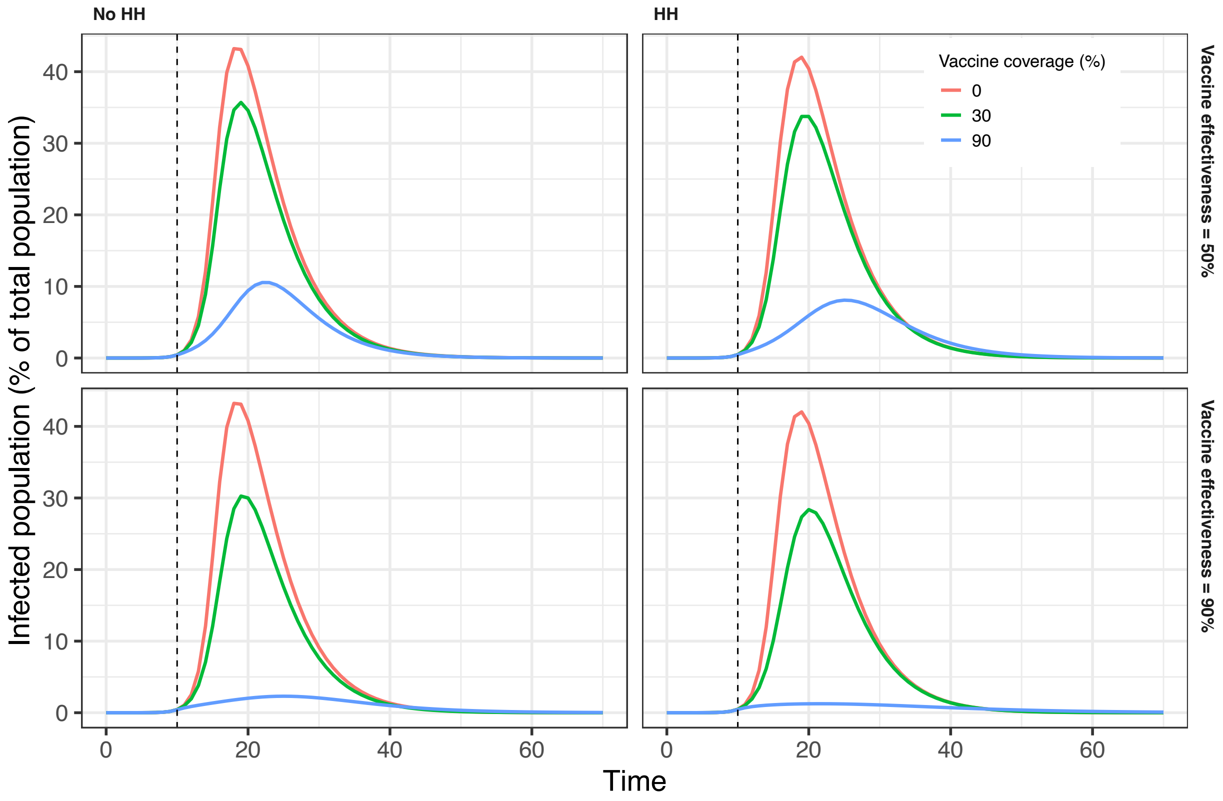


Figure A.2: Epidemic curves under different vaccination coverage and effectiveness for E=1 and I=1

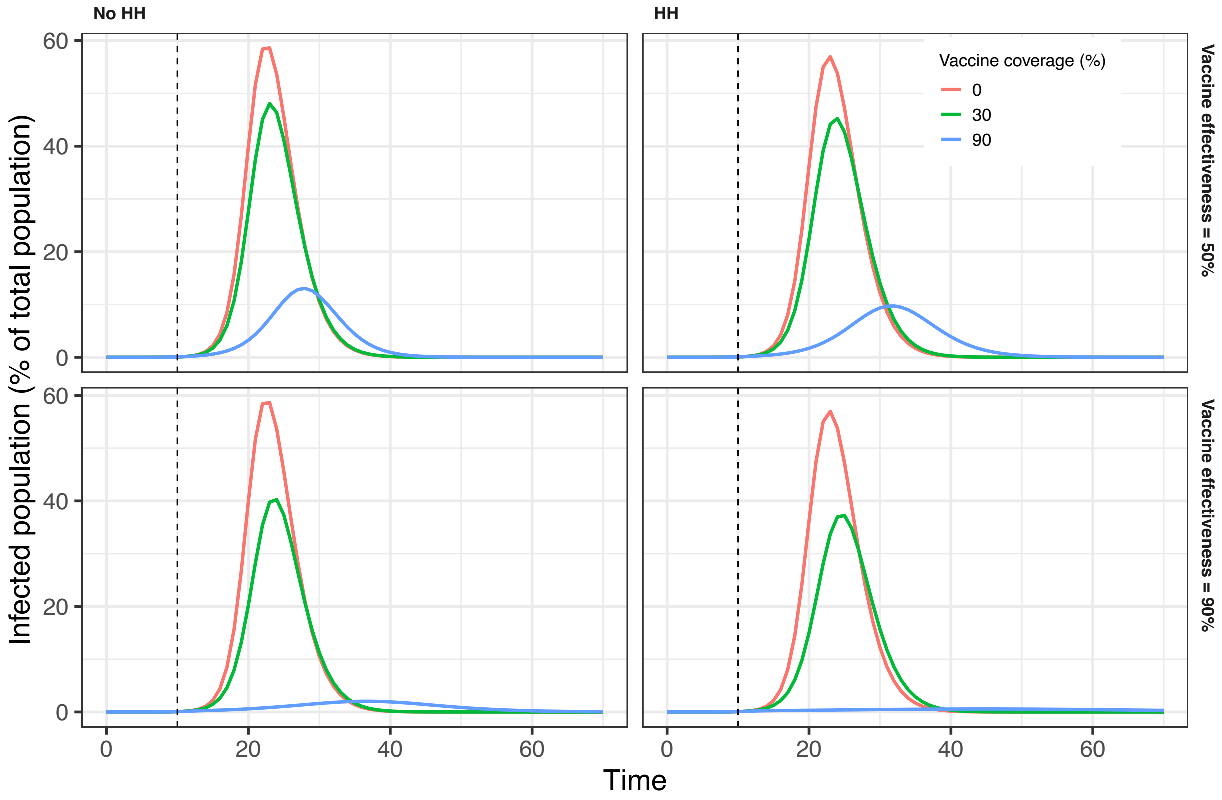


Figure A.3: Epidemic curves under different vaccination coverage and effectiveness for E=3 and I=3.

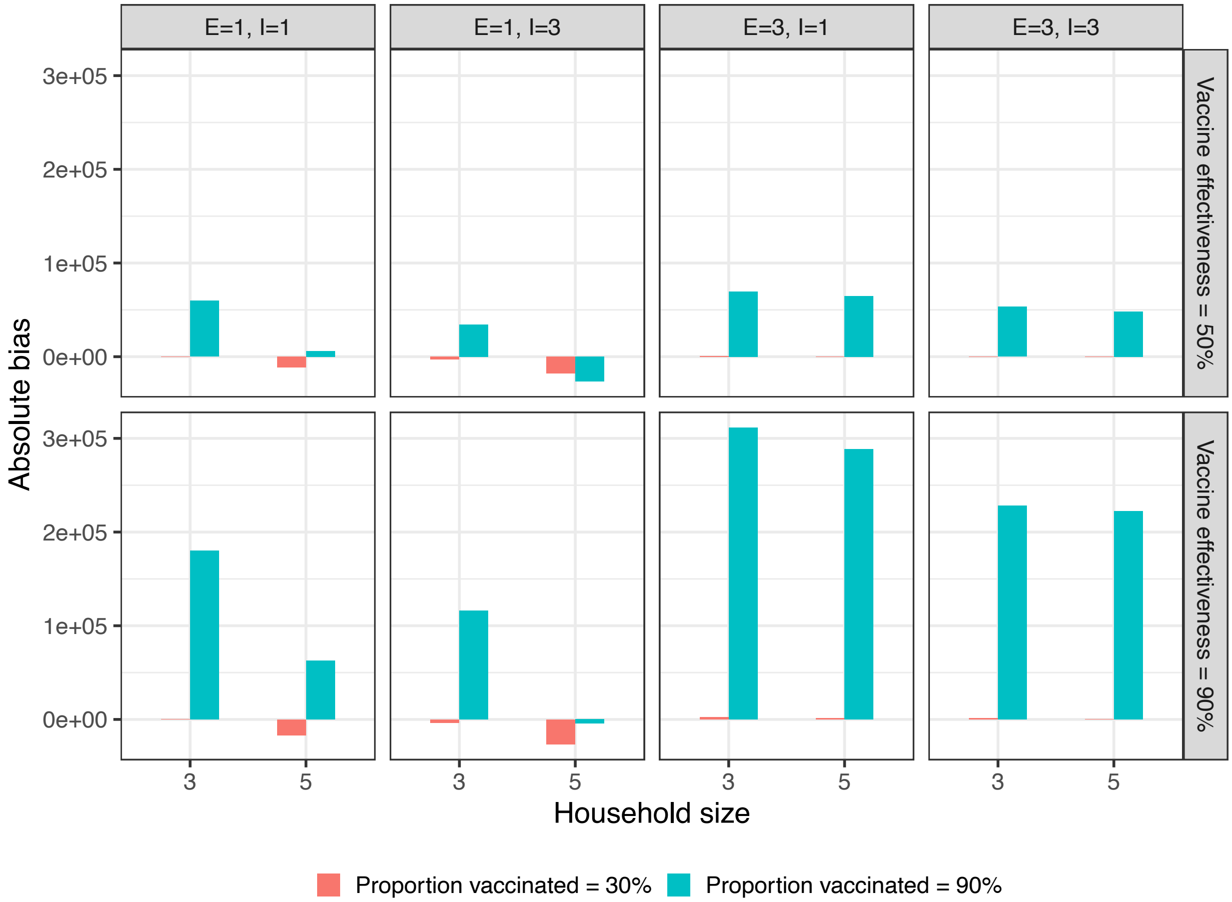


Figure A.4: Vaccine effectiveness absolute bias on epidemic size.

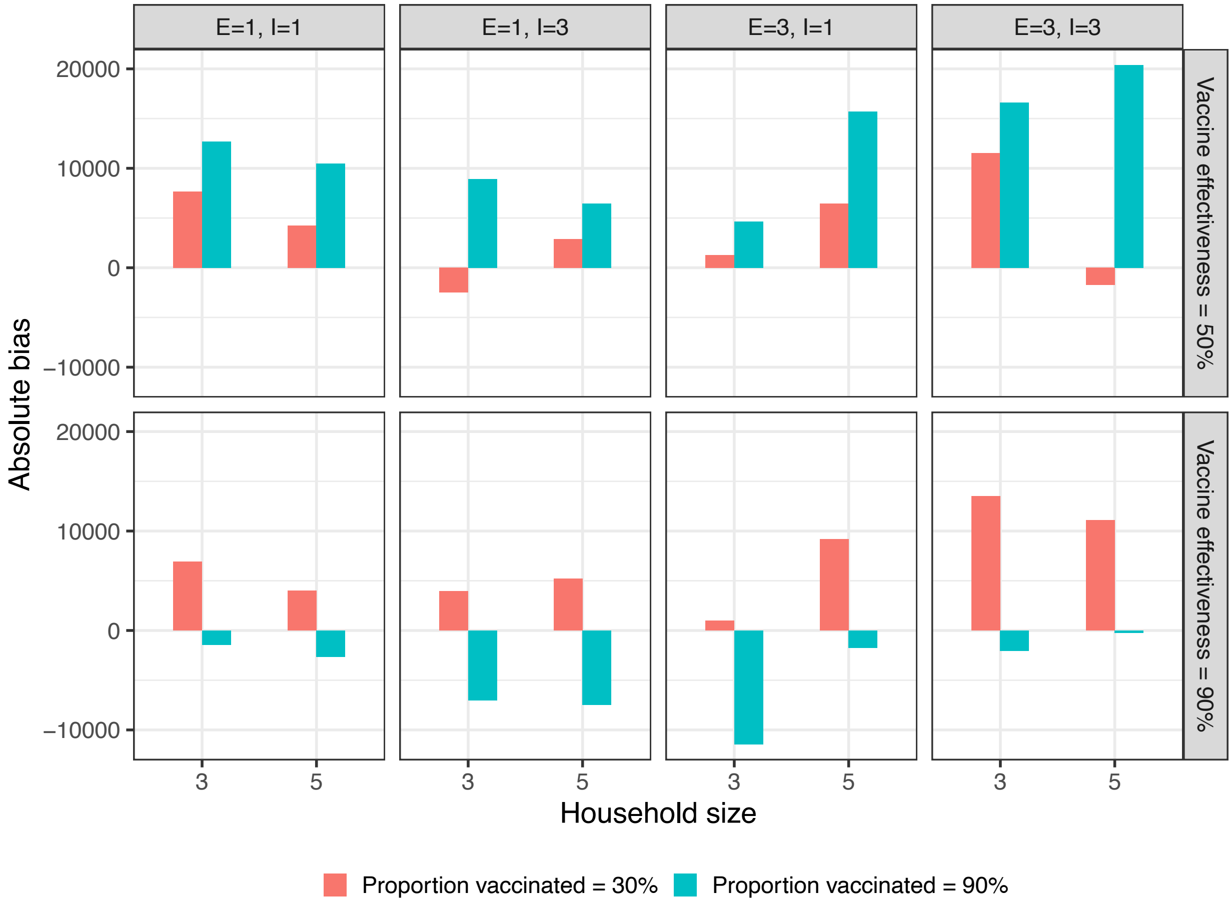


Figure A.5: Vaccine effectiveness absolute bias on peak size.

Table A10: Meta-regression estimates on absolute bias of effects of vaccination coverage = 30%, effectiveness = 50%, and $\omega$ = 0.

| | Peak time (days) | | Peak size (people) | | Epidemic size (people) | |
|---|---|---|---|---|---|---|
| Household size 3 (HH3) | -0.3 | -0.3 | -1,158.0 | 226.1 | -5,154.9 | -1,830.6 |
| Household size 5 (HH5) | 0.1 | 0.03 | 1,151.1 | -1,617.2 | -2,227.0** | -8,875.6*** |
| E | -0.1 | -0.1 | 1,727.8*** | 1,129.9 | 4,017.3*** | 2,897.9*** |
| I | 0.1 | 0.1 | 289.5 | 195.3 | -133.7 | -676.4 |
| $\tau$ | 1.1 | 1.1 | -17,791.8* | -17,791.8* | -11,735.8 | -11,735.8 |
| $\beta$ | -0.6 | -0.6 | 12,162.3 | 12,162.3 | -1,958.2 | -1,958.2 |
| E*HH5 | | 0.1 | | 1,195.8 | | 2,238.9** |
| I*HH5 | | -0.04 | | 188.4 | | 1,085.4 |
| HH3 + HH5 | -0.2 | -0.2 | -7.0 | -7.0 | -7,381.9 | -7,381.9 |

*Note:* *p<0.1; **p<0.05; ***p<0.01
Absolute bias is equal to effect without HH structure minus effect with structure.
Interventions increase time to peak; Positive bias means the model without HH structure yields a larger effect.
Interventions decrease the size of the peak; Negative bias means the model without HH structure yields a larger effect.
Interventions decrease epidemic size; Negative bias means the model without HH structure yields a larger effect.
HH3 describes how much the intervention effect differs if the household structure is excluded from an otherwise similar epidemic where the true household size is 3.
HH5 describes the incremental difference in the intervention effect if household structure is excluded from an otherwise similar epidemic where the true household size is 5 instead of 3.
For the models, the total effect of exclusion of household structure involves interactions with other terms.
For example, E*HH5 describes how the incremental difference depends upon the number of exposed compartments when the true household size is 5 instead of 3.
We estimated the magnitude of the linear combination of the relevant coefficients and tested their significance.
E: Number of exposed compartments; I: Number of infectious compartments; $\tau$: Household transmission rate; $\beta$: Community transmission rate; $\omega$: Waning immunity rate.

Table A11: Meta-regression estimates on absolute bias of effects of vaccination coverage = 90%, effectiveness = 50%, and $\omega$ = 0.

| | Peak time (days) | | Peak size (people) | | Epidemic size (people) | |
|---|---|---|---|---|---|---|
| Household size 3 (HH3) | -7.0*** | -7.4*** | 9,131.4 | 16,052.6*** | 127,412.7*** | 139,950.8*** |
| Household size 5 (HH5) | 0.7*** | 1.6*** | 4,997.4*** | -8,845.0*** | -8,983.3** | -34,059.6** |
| E | -1.1*** | -0.9*** | 3,699.5*** | 860.8 | 15,971.5*** | 11,881.1*** |
| I | 0.5*** | 0.5*** | 1,099.2* | 477.2 | -5,195.0** | -7,373.7** |
| $\tau$ | 16.7*** | 16.7*** | -50,482.7*** | -50,482.7*** | -388,528.5*** | -388,528.5*** |
| $\beta$ | 1.7 | 1.7 | 10,075.4 | 10,075.4 | -5,757.5 | -5,757.5 |
| E*HH5 | | -0.3 | | 5,677.3*** | | 8,180.8* |
| I*HH5 | | -0.1 | | 1,243.9 | | 4,357.4 |
| HH3 + HH5 | -6.3*** | -6.3*** | 14,128.8** | 14,128.8*** | 118,429.4*** | 118,429.4*** |

*Note:* *p<0.1; **p<0.05; ***p<0.01
Absolute bias is equal to effect without HH structure minus effect with structure.
Interventions increase time to peak; Positive bias means the model without HH structure yields a larger effect.
Interventions decrease the size of the peak; Negative bias means the model without HH structure yields a larger effect.
Interventions decrease epidemic size; Negative bias means the model without HH structure yields a larger effect.
HH3 describes how much the intervention effect differs if the household structure is excluded from an otherwise similar epidemic where the true household size is 3.
HH5 describes the incremental difference in the intervention effect if the household structure is excluded from an otherwise similar epidemic where the true household size is 5 instead of 3.
For the models, the total effect of exclusion of household structure involves interactions with other terms.
For example, E*HH5 describes how the incremental difference depends upon the number of exposed compartments when the true household size is 5 instead of 3.
We estimated the magnitude of the linear combination of the relevant coefficients and tested their significance.
E: Number of exposed compartments; I: Number of infectious compartments; $\tau$: Household transmission rate; $\beta$: Community transmission rate; $\omega$: Waning immunity rate.

Table A12: Meta-regression estimates on absolute bias of effects of vaccination coverage = 30%, effectiveness = 90%, and $\omega$ = 0.

| | Peak time (days) | | Peak size (people) | | Epidemic size (people) | |
|---|---|---|---|---|---|---|
| Household size 3 (HH3) | -0.5 | -0.5 | 6,583.6 | 9,700.6$^{**}$ | -5,863.9 | -952.6 |
| Household size 5 (HH5) | 0.1 | 0.1 | 2,182.0$^{***}$ | -4,051.8$^{*}$ | -3,165.2$^{**}$ | -12,988.0$^{**}$ |
| E | -0.2$^{**}$ | -0.2$^{*}$ | 2,446.2$^{***}$ | 1,272.9$^{**}$ | 5,965.8$^{***}$ | 4,428.8$^{***}$ |
| I | 0.04 | 0.1 | 691.6 | 306.4 | -235.2 | -1,153.9 |
| $\tau$ | 3.6$^{***}$ | 3.6$^{***}$ | -35,010.7$^{***}$ | -35,010.7$^{***}$ | -22,713.3 | -22,713.3 |
| $\beta$ | -1.9 | -1.9 | 5,267.3 | 5,267.3 | -2,124.6 | -2,124.6 |
| E*HH5 | | 0.1 | | 2,346.5$^{***}$ | | 3,073.9$^{*}$ |
| I*HH5 | | -0.1 | | 770.4 | | 1,837.5 |
| HH3 + HH5 | -0.3 | -0.3 | 8,765.7$^{**}$ | 8,765.7$^{**}$ | -9,029.2 | -9,029.2 |

*Note:* $^{*}$p<0.1; $^{**}$p<0.05; $^{***}$p<0.01
Absolute bias is equal to effect without HH structure minus effect with structure.
Interventions increase time to peak; Positive bias means the model without HH structure yields a larger effect.
Interventions decrease the size of the peak; Negative bias means the model without HH structure yields a larger effect.
Interventions decrease epidemic size; Negative bias means the model without HH structure yields a larger effect.
HH3 describes how much the intervention effect differs if the household structure is excluded from an otherwise similar epidemic where the true household size is 3.
HH5 describes the incremental difference in the intervention effect if the household structure is excluded from an otherwise similar epidemic where the true household size is 5 instead of 3.
For the models, the total effect of exclusion of household structure involves interactions with other terms.
For example, E*HH5 describes how the incremental difference depends upon the number of exposed compartments when the true household size is 5 instead of 3.
We estimated the magnitude of the linear combination of the relevant coefficients and tested their significance.
E: Number of exposed compartments; I: Number of infectious compartments; $\tau$: Household transmission rate; $\beta$: Community transmission rate; $\omega$: Waning immunity rate.

Table A13: Meta-regression estimates on absolute bias of effects of vaccination coverage = 90%, effectiveness = 90%, and $\omega$ = 0.

| | Peak time (days) | | Peak size (people) | | Epidemic size (people) | |
|---|---|---|---|---|---|---|
| Household size 3 (HH3) | -3.4 | -1.3 | -22,018.4*** | -17,024.5*** | 449,115.2*** | 481,855.8*** |
| Household size 5 (HH5) | 2.1*** | -2.1 | 4,803.4*** | -5,184.5* | -25,386.6*** | -90,867.8*** |
| E | -3.1*** | -4.6*** | 1,650.9*** | -224.7 | 67,071.4*** | 54,368.6*** |
| I | 0.7* | 1.1** | 406.0 | -215.3 | -22,370.2*** | -26,037.7*** |
| $\tau$ | 10.3* | 10.3** | 35,223.4*** | 35,223.4*** | -1,277,971.0*** | -1,277,971.0*** |
| $\beta$ | 5.3 | 5.3 | 7,907.2 | 7,907.2 | -34,507.9 | -34,507.9 |
| E*HH5 | | 3.0*** | | 3,751.3*** | | 25,405.6** |
| I*HH5 | | -0.8 | | 1,242.6 | | 7,335.1 |
| HH3 + HH5 | -1.3 | -1.3 | -17,215.0*** | -17,215.0*** | 423,728.6*** | 423,728.6*** |

*Note:* *p<0.1; **p<0.05; ***p<0.01
Absolute bias is equal to effect without HH structure minus effect with structure.
Interventions increase time to peak; Positive bias means the model without HH structure yields a larger effect.
Interventions decrease the size of the peak; Negative bias means the model without HH structure yields a larger effect.
Interventions decrease epidemic size; Negative bias means the model without HH structure yields a larger effect.
HH3 describes how much the intervention effect differs if the household structure is excluded from an otherwise similar epidemic where the true household size is 3.
HH5 describes the incremental difference in the intervention effect if the household structure is excluded from an otherwise similar epidemic where the true household size is 5 instead of 3.
For the models, the total effect of exclusion of household structure involves interactions with other terms.
For example, E*HH5 describes how the incremental difference depends upon the number of exposed compartments when the true household size is 5 instead of 3.
We estimated the magnitude of the linear combination of the relevant coefficients and tested their significance.
E: Number of exposed compartments; I: Number of infectious compartments; $\tau$: Household transmission rate; $\beta$: Community transmission rate; $\omega$: Waning immunity rate.
